# Long-term COVID-19 booster effectiveness by infection history and clinical vulnerability and immune imprinting

**DOI:** 10.1101/2022.11.14.22282103

**Authors:** Hiam Chemaitelly, Houssein H. Ayoub, Patrick Tang, Peter Coyle, Hadi M. Yassine, Asmaa A. Al Thani, Hebah A. Al-Khatib, Mohammad R. Hasan, Zaina Al-Kanaani, Einas Al-Kuwari, Andrew Jeremijenko, Anvar Hassan Kaleeckal, Ali Nizar Latif, Riyazuddin Mohammad Shaik, Hanan F. Abdul-Rahim, Gheyath K. Nasrallah, Mohamed Ghaith Al-Kuwari, Adeel A. Butt, Hamad Eid Al-Romaihi, Mohamed H. Al-Thani, Abdullatif Al-Khal, Roberto Bertollini, Jeremy Samuel Faust, Laith J. Abu-Raddad

**Author notes:** Correspondence to Dr. Hiam Chemaitelly, or Professor Laith J. Abu-Raddad,.

## Abstract

**Background:** Long-term effectiveness of COVID-19 mRNA boosters in populations with different prior infection histories and clinical vulnerability profiles is inadequately understood.

**Methods:** A national, matched, retrospective, target trial cohort study was conducted in Qatar to investigate effectiveness of a third mRNA (booster) dose, relative to a primary series of two doses, against SARS-CoV-2 omicron infection and against severe COVID-19. Associations were estimated using Cox proportional-hazards regression models.

**Results:** Booster effectiveness relative to primary series was 41.1% (95% CI: 40.0-42.1%) against infection and 80.5% (95% CI: 55.7-91.4%) against severe, critical, or fatal COVID-19, over one-year follow-up after the booster. Among persons clinically vulnerable to severe COVID-19, effectiveness was 49.7% (95% CI: 47.8-51.6%) against infection and 84.2% (95% CI: 58.8-93.9%) against severe, critical, or fatal COVID-19. Effectiveness against infection was highest at 57.1% (95% CI: 55.9-58.3%) in the first month after the booster but waned thereafter and was modest at only 14.4% (95% CI: 7.3-20.9%) by the sixth month. In the seventh month and thereafter, coincident with BA.4/BA.5 and BA.2.75* subvariant incidence, effectiveness was progressively negative reaching -20.3% (95% CI: -55.0-29.0%) after one year of follow-up. Similar levels and patterns of protection were observed irrespective of prior infection status, clinical vulnerability, or type of vaccine (BNT162b2 versus mRNA-1273).

**Conclusions:** Boosters reduced infection and severe COVID-19, particularly among those clinically vulnerable to severe COVID-19. However, protection against infection waned after the booster, and eventually suggested an imprinting effect of compromised protection relative to the primary series. However, imprinting effects are unlikely to negate the overall public health value of booster vaccinations.

## Introduction

With waning of vaccine and natural infection protection against the severe acute respiratory syndrome coronavirus 2 (SARS-CoV-2) infection and against severe coronavirus disease 2019 (COVID-19),^1-4^ repeat booster vaccination may sustain immune protection against infection and disease.^5,6^ However, the global population carries heterogenous immune histories due to varying exposures to infection from different viral variants, and vaccination.^7^ Booster effectiveness may vary by prior infection and vaccination history, prior variant exposure, and by age and clinical vulnerability to severe COVID-19. Immune imprinting, a phenomenon in which the specific sequence of immunological events (due to infection and/or vaccination) can enhance or compromise a person’s future immune protection, could affect the utility of booster vaccination.^7-10^ The optimal public health impact of boosters may not be achieved through a “one prescription fits all” approach.

We investigated the long-term real-world effectiveness of booster (third dose) vaccination against SARS-CoV-2 infection and against severe,^11^ critical,^11^ or fatal^12^ COVID-19, relative to that of primary-series (two-dose) vaccination, in persons with different immune histories and different clinical vulnerability to infection, over a follow-up duration of one year.

## Methods

### Study population and data sources

This study was conducted on the population of Qatar including data between January 5, 2021, earliest record of second dose vaccination, and October 12, 2022. It analyzed the national, federated databases for COVID-19 laboratory testing, vaccination, hospitalization, and death, retrieved from the integrated, nationwide, digital-health information platform (Section S1 of the Supplementary Appendix). Databases include all SARS-CoV-2-related data with no missing information since the onset of the pandemic, including all polymerase chain reaction (PCR) tests, and from January 5, 2022 onward, all rapid antigen tests conducted at healthcare facilities (Section S2). SARS-CoV-2 testing in Qatar is done at mass scale, mostly for routine reasons.^2,13^ Most infections are diagnosed not because of symptoms, but because of routine testing.^2,13^ Qatar launched its COVID-19 vaccination program in December of 2020 using BNT162b2 and mRNA-1273.^14^ Detailed descriptions of Qatar’s population and of the national databases have been reported previously.^2,5,13,15,16^

### Study design and cohorts

We conducted a matched, retrospective, cohort study that emulated a randomized “target” trial.^5,17^ Incidence of breakthrough infection and associated severe, critical, or fatal COVID-19 were compared in the national cohort of persons who received a third (booster) vaccine dose (designated the three-dose cohort) to that in the national cohort of persons who received the primary-series (designated the two-dose cohort). Incidence of infection was defined as the first PCR-positive or rapid-antigen-positive test after the start of follow-up, regardless of symptoms. Infection severity classification followed World Health Organization guidelines for COVID-19 case severity (acute-care hospitalizations),^11^ criticality (intensive-care-unit hospitalizations),^11^ and fatality^12^ (Section S3).

Incidence was also compared for subgroups including persons with no prior infection, prior infection with either pre-omicron or omicron (B.1.1.529) viruses, and prior infections with both viruses. Prior infections were classified as pre-omicron if they occurred before December 19, 2021, the date of onset of the omicron wave in Qatar,^18^ and as omicron otherwise.

Incidence was also compared in the subgroup of persons who are less clinically vulnerable to severe COVID-19, defined as persons who are below 50 years of age and with one or no coexisting conditions,^19^ and in the subgroup of persons who are more clinically vulnerable to severe COVID-19, defined as persons who are ≥50 years of age, or who are <50 years of age but with ≥2 coexisting conditions (Section S1).^19^ Incidence was also compared between those vaccinated with BNT162b2 and with mRNA-1273.

### Cohorts matching and follow-up

Cohorts were matched exactly one-to-one by sex, 10-year age group, nationality, number of coexisting conditions (0, 1, 2, 3, 4, 5, or ≥6 coexisting conditions), vaccine type (BNT162b2 or mRNA-1273), and prior infection status (no prior infection, or prior infection with either pre-omicron or omicron viruses, or prior infections with both viruses) to balance observed confounders between exposure groups that are related to infection risk in Qatar.^15,20-23^ Matching by these factors was previously shown to provide adequate control of differences in exposure risk in Qatar.^2,14,24-26^

To control for time since second-dose vaccination, matching was also done by calendar week of the second dose (i.e., matched pairs had to have second doses in the same calendar week). Persons receiving their third vaccine dose in a specific calendar week in the three-dose cohort were additionally matched to persons in the two-dose cohort with records for SARS-CoV-2 testing in that same calendar week, ensuring that matched pairs were present in Qatar in the same period.

Persons were eligible for inclusion in the three-dose cohort if they received three vaccine doses with the same mRNA vaccine and had no record for a SARS-CoV-2-positive test within 90 days before the start of follow-up. The latter exclusion criterion ensured that infections after start of follow-up were incident infections and not prolonged SARS-CoV-2-positivity of earlier infections.^18,27,28^ Persons were eligible for inclusion in the two-dose cohort if they received two doses of the same mRNA vaccine. Persons receiving the pediatric BNT162b2 vaccine were excluded from both cohorts.

Matching was performed iteratively such that persons in the two-dose cohort were alive and had no record before the start date of follow-up for a third dose or for a SARS-CoV-2-positive test within 90 days. Persons in the two-dose cohort contributed follow-up time before receiving the third dose (and matched to three-dose-vaccinated persons), and subsequently contributed follow-up time in the three-dose cohort, if they received a third dose (and matched to two-dose-vaccinated persons). Matching was iterated with as many replications as needed until exhaustion (i.e., no more matched pairs could be identified).

As in previous studies,^5,29^ to ensure time for sufficient immunogenicity, both members of each matched pair were followed starting 7 days after the calendar date in which the person in the three-dose cohort received the third dose. For exchangeability,^5,29^ both members of each matched pair were censored at earliest occurrence of the person in the three-dose cohort receiving the fourth dose or the person in the two-dose cohort receiving the third dose. Accordingly, individuals were followed up until the first of any of the following events: a documented SARS-CoV-2 infection, or fourth-dose vaccination for persons in the three-dose cohort (with matched-pair censoring), or third-dose vaccination for persons in the two-dose cohort (with matched-pair censoring), or death, or end of study censoring (October 12, 2022).

### Oversight

The institutional review boards at Hamad Medical Corporation and Weill Cornell Medicine– Qatar approved this retrospective study with a waiver of informed consent. The study was reported according to the Strengthening the Reporting of Observational Studies in Epidemiology (STROBE) guidelines (Table S1). The authors vouch for the accuracy and completeness of the data and for the fidelity of the study to the protocol. Data used in this study are the property of the Ministry of Public Health of Qatar and were provided to the researchers through a restricted-access agreement for preservation of confidentiality of patient data. The funders had no role in the study design; the collection, analysis, or interpretation of the data; or the writing of the manuscript.

### Statistical analysis

Eligible and matched cohorts were described using frequency distributions and measures of central tendency and were compared using standardized mean differences (SMDs). An SMD of ≤0.1 indicated adequate matching.^30^ Cumulative incidence of infection (defined as proportion of persons at risk, whose primary endpoint during follow-up was an infection) was estimated using the Kaplan-Meier estimator method. Incidence rate of infection in each cohort, defined as number of identified infections divided by number of person-weeks contributed by all individuals in the cohort, was estimated, with the corresponding 95% confidence interval (CI) using a Poisson log-likelihood regression model with the Stata 17.0 *stptime* command.

Hazard ratios (HR), comparing incidence of infection in the cohorts and corresponding 95% CIs, were calculated using Cox regression adjusted for the matching factors with the Stata 17.0 *stcox* command. Sensitivity analysis further adjusting the HR for differences in testing frequency between cohorts was conducted. Schoenfeld residuals and log-log plots for survival curves were used to test the proportional-hazards assumption. CIs were not adjusted for multiplicity; thus, they should not be used to infer definitive differences between groups. Interactions were not considered. Vaccine effectiveness was estimated as 1-adjusted hazard ratio (aHR) if the aHR was <1, and as 1/aHR-1 if the aHR was ≥1. The latter was to ensure symmetric scale for both negative and positive effectiveness, ranging from -100%-100%.

Additional analyses were conducted to investigate waning of booster protection over time. aHRs were estimated by month since the start of follow-up using separate Cox regressions with “failures” restricted to specific months. Statistical analyses were performed using Stata/SE version 17.0 (Stata Corporation, College Station, TX, USA).

## Results

### Overall booster effectiveness

Figure S1 shows the study population selection process. Table 1 describes baseline characteristics of the full and matched cohorts. Matched cohorts each included 304,091 persons.

**Table 1:** Baseline characteristics of eligible and matched cohorts in the study investigating effectiveness of mRNA booster (third dose) vaccination relative to that of primary-series (two-dose) vaccination.

| Characteristics | Full eligible cohorts |  |  | Matched cohorts<sup>*</sup> |  |  |
| --- | --- | --- | --- | --- | --- | --- |
|  | Three-dose cohort | Two-dose cohort | SMD<sup>‡</sup> | Three-dose cohort | Two-dose cohort | SMD<sup>‡</sup> |
|  | N=658,947 | N=2,228,686 |  | N=304,091 | N=304,091 |  |
| Median age (IQR)—years | 39 (32-47) | 36 (30-44) | 0.23<sup>‡</sup> | 37 (31-44) | 37 (31-44) | 0.01<sup>‡</sup> |
| Age—years |  |  |  |  |  |  |
| 0-9 years | 1 (<0.01) | 11 (<0.01) |  | -- | -- |  |
| 10-19 years | 39,663 (6.0) | 132,021 (5.9) |  | 11,909 (3.9) | 11,909 (3.9) |  |
| 20-29 years | 80,310 (12.2) | 419,633 (18.8) |  | 44,296 (14.6) | 44,296 (14.6) |  |
| 30-39 years | 227,636 (34.6) | 840,553 (37.7) | 0.26 | 127,304 (41.9) | 127,304 (41.9) | 0.00 |
| 40-49 years | 172,513 (26.2) | 530,743 (23.8) |  | 80,707 (26.5) | 80,707 (26.5) |  |
| 50-59 years | 93,428 (14.2) | 219,088 (9.8) |  | 31,225 (10.3) | 31,225 (10.3) |  |
| 60-69 years | 35,866 (5.4) | 67,905 (3.1) |  | 7,473 (2.5) | 7,473 (2.5) |  |
| 70+ years | 9,530 (1.5) | 18,732 (0.8) |  | 1,177 (0.4) | 1,177 (0.4) |  |
| Sex |  |  |  |  |  |  |
| Male | 432,830 (65.7) | 1,644,730 (73.8) | 0.18 | 225,155 (74.0) | 225,155 (74.0) | 0.00 |
| Female | 226,117 (34.3) | 583,956 (26.2) |  | 78,936 (26.0) | 78,936 (26.0) |  |
| Nationality<sup>§</sup> |  |  |  |  |  |  |
| Bangladeshi | 57,949 (8.8) | 312,144 (14.0) |  | 33,199 (10.9) | 33,199 (10.9) |  |
| Egyptian | 54,168 (8.2) | 110,227 (5.0) |  | 17,473 (5.8) | 17,473 (5.8) |  |
| Filipino | 89,736 (13.6) | 208,781 (9.4) |  | 34,734 (11.4) | 34,734 (11.4) |  |
| Indian | 207,941 (31.6) | 549,301 (24.7) |  | 110,465 (36.3) | 110,465 (36.3) |  |
| Nepalese | 26,055 (4.0) | 239,077 (10.7) | 0.39 | 18,474 (6.1) | 18,474 (6.1) | 0.00 |
| Pakistani | 31,132 (4.7) | 106,388 (4.8) |  | 12,946 (4.3) | 12,946 (4.3) |  |
| Qatari | 39,301 (6.0) | 199,432 (9.0) |  | 29,820 (9.8) | 29,820 (9.8) |  |
| Sri Lankan | 19,069 (2.9) | 77,844 (3.5) |  | 9,490 (3.1) | 9,490 (3.1) |  |
| Sudanese | 11,777 (1.8) | 46,267 (2.1) |  | 3,800 (1.3) | 3,800 (1.3) |  |
| Other nationalities<sup>¶</sup> | 121,819 (18.5) | 379,225 (17.0) |  | 33,690 (11.1) | 33,690 (11.1) |  |
| Number of coexisting conditions |  |  |  |  |  |  |
| None | 494,154 (75.0) | 1,857,593 (83.4) |  | 265,210 (87.2) | 265,210 (87.2) |  |
| 1 | 73,796 (11.2) | 189,741 (8.5) |  | 21,078 (6.9) | 21,078 (6.9) |  |
| 2 | 42,121 (6.4) | 89,898 (4.0) |  | 9,388 (3.1) | 9,388 (3.1) |  |
| 3 | 21,633 (3.3) | 41,405 (1.9) | 0.22 | 3,905 (1.3) | 3,905 (1.3) | 0.00 |
| 4 | 12,952 (2.0) | 23,532 (1.1) |  | 2,159 (0.7) | 2,159 (0.7) |  |
| 5 | 7,338 (1.1) | 13,409 (0.6) |  | 1,083 (0.4) | 1,083 (0.4) |  |
| 6+ | 6,953 (1.1) | 13,108 (0.6) |  | 1,268 (0.4) | 1,268 (0.4) |  |
| Vaccine type |  |  |  |  |  |  |
| BNT162b2 | 429,788 (65.2) | 1,322,503 (59.3) | 0.12 | 184,737 (60.8) | 184,737 (60.8) | 0.00 |
| mRNA-1273 | 229,159 (34.8) | 906,183 (40.7) |  | 119,354 (39.2) | 119,354 (39.2) |  |
| Prior infection status<sup>**</sup> |  |  |  |  |  |  |
| No prior infection | 549,277 (83.4) | -- |  | 267,107 (87.8) | 267,107 (87.8) |  |
| Pre-Omicron | 90,307 (13.7) | -- |  | 30,755 (10.1) | 30,755 (10.1) |  |
| Omicron | 17,500 (2.7) | -- | -- | 6,023 (2.0) | 6,023 (2.0) | 0.00 |
| Pre-Omicron & Omicron | 1,863 (0.3) | -- |  | 206 (0.1) | 206 (0.1) |  |

IQR denotes interquartile range, SARS-CoV-2 severe acute respiratory syndrome coronavirus 2, and SMD standardized mean difference.
<sup>\*</sup>Cohorts were matched exactly one-to-one by sex, 10-year age group, nationality, number of coexisting conditions, vaccine type, prior infection status, and calendar week of the second vaccine dose. Persons who received their third vaccine dose in a specific calendar week in the three-dose cohort were additionally matched to persons who had a record for a SARS-CoV-2 test in that same calendar week in the two-dose cohort, to ensure that matched pairs had presence in Qatar over the same time period.
<sup>‡</sup>SMD is the difference in the mean of a covariate between groups divided by the pooled standard deviation. An SMD ≤0.1 indicates adequate matching.
<sup>‡</sup>SMD is for the mean difference between groups divided by the pooled standard deviation.
<sup>§</sup>Nationalities were chosen to represent the most populous groups in Qatar.
<sup>¶</sup>These comprise up to 173 other nationalities in the unmatched cohorts, and 106 other nationalities in the matched cohorts.
<sup>\*\*</sup>Ascertained at the start of follow-up. Accordingly, distribution is not available for the unmatched two-dose cohort, as the start of follow-up for each person in the two-dose cohort is determined by that of their match in the three-dose cohort (7 days after the third dose) after the matching is performed.

For the matched three-dose cohort, median date of first dose was April 15, 2021, of second dose was May 12, 2021, and of third dose was January 16, 2022. Median duration between first and second doses was 22 days (interquartile range (IQR), 21-28 days) and between second and third doses was 249 days (IQR, 221-282 days). For the matched two-dose cohort, median date of first dose was April 15, 2021, and of second dose was May 12, 2021. Median duration between first and second doses was 22 days (IQR, 21-28 days).

Median duration of follow-up was 203 days (IQR, 59-262 days) for the three-dose cohort and 190 days (IQR, 45-256 days) for the two-dose cohort (Figure 1). During follow-up, 20,528 infections were recorded in the three-dose cohort, of which 7 progressed to severe, but none to critical or fatal COVID-19 (Figure S1). Meanwhile, 30,771 infections were recorded in the two-dose cohort, of which 25 progressed to severe, 3 to critical, and 3 to fatal COVID-19.

**Figure 1:**
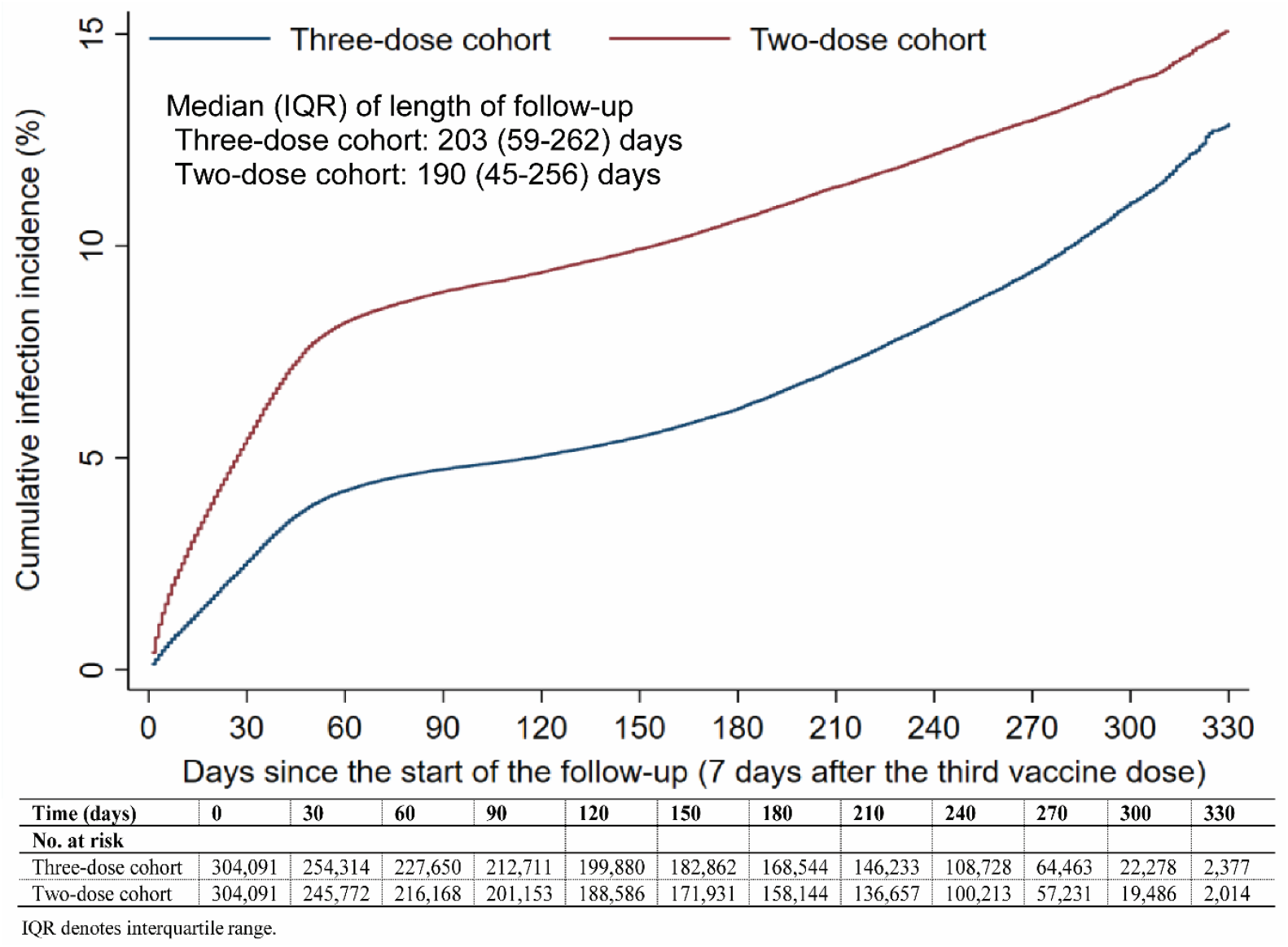
Cumulative incidence of SARS-CoV-2 infection in the matched three-dose and two-dose vaccination cohorts.

Cumulative incidence of infection was 12.9% (95% CI: 12.5-13.3%) in the three-dose cohort and 15.1% (95% CI: 14.7-15.4%) in the two-dose cohort, 330 days after the start of follow-up (Figure 1). Incidence during follow-up was dominated by omicron subvariants including first a large BA.1/BA.2 wave,^31^ and subsequently BA.4/BA.5^32^ and BA.2.75* (predominantly BA.2.75.2)^33^ waves. A small proportion of the cohorts experienced low B.1.617.2 (delta) incidence, but only for a very short duration of follow-up.^5^

aHR comparing incidence of infection in the three-dose cohort to the two-dose cohort was 0.59 (95% CI: 0.58-0.60; Table 2). Booster effectiveness against infection was 41.1% (95% CI: 40.0-42.1%). aHR for incidence of severe, critical, or fatal COVID-19 was 0.19 (95% CI: 0.09-0.44). Booster effectiveness against severe, critical, or fatal COVID-19 was 80.5% (95% CI: 55.7-91.4%).

**Table 2:**
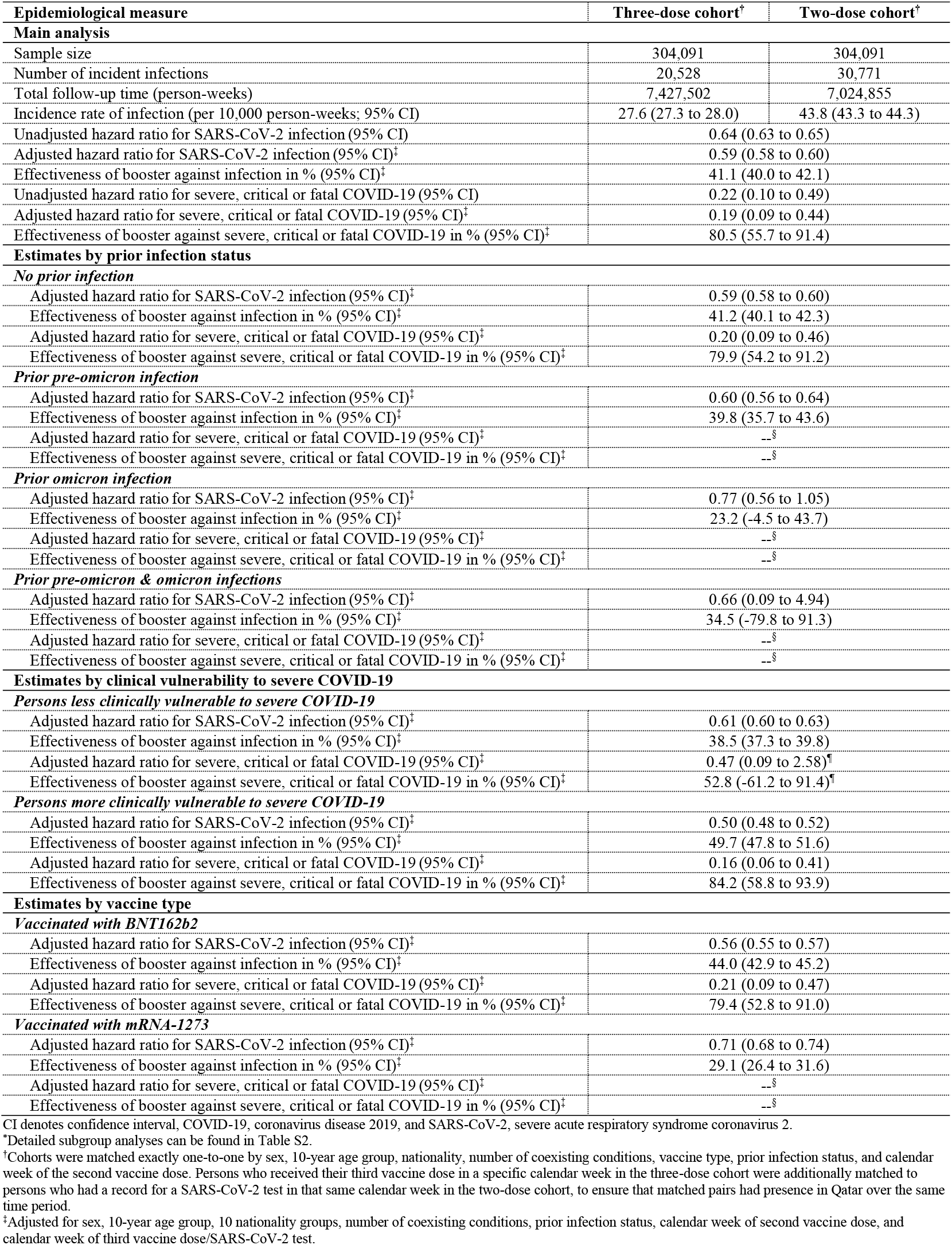
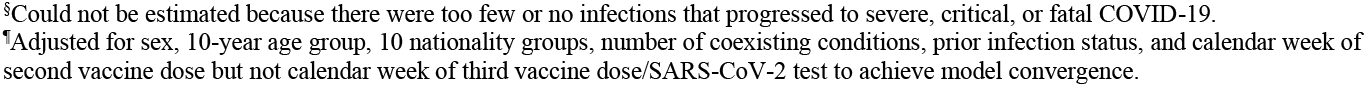
Hazard ratios for incidence of SARS-CoV-2 infection and severe, critical or fatal COVID-19 in the three-dose cohort versus the two-dose cohort.^*^

| Epidemiological measure | Three-dose cohort<sup>†</sup> | Two-dose cohort<sup>‡</sup> |
| --- | --- | --- |
| **Main analysis** |  |  |
| Sample size | 304,091 | 304,091 |
| Number of incident infections | 20,528 | 30,771 |
| Total follow-up time (person-weeks) | 7,427,502 | 7,024,855 |
| Incidence rate of infection (per 10,000 person-weeks; 95% CI) | 27.6 (27.3 to 28.0) | 43.8 (43.3 to 44.3) |
| Unadjusted hazard ratio for SARS-CoV-2 infection (95% CI) | 0.64 (0.63 to 0.65) |  |
| Adjusted hazard ratio for SARS-CoV-2 infection (95% CI)<sup>‡</sup> | 0.59 (0.58 to 0.60) |  |
| Effectiveness of booster against infection in % (95% CI)<sup>‡</sup> | 41.1 (40.0 to 42.1) |  |
| Unadjusted hazard ratio for severe, critical or fatal COVID-19 (95% CI) | 0.22 (0.10 to 0.49) |  |
| Adjusted hazard ratio for severe, critical or fatal COVID-19 (95% CI)<sup>‡</sup> | 0.19 (0.09 to 0.44) |  |
| Effectiveness of booster against severe, critical or fatal COVID-19 in % (95% CI)<sup>‡</sup> | 80.5 (55.7 to 91.4) |  |
| **Estimates by prior infection status** |  |  |
| *No prior infection* |  |  |
| Adjusted hazard ratio for SARS-CoV-2 infection (95% CI)<sup>‡</sup> | 0.59 (0.58 to 0.60) |  |
| Effectiveness of booster against infection in % (95% CI)<sup>‡</sup> | 41.2 (40.1 to 42.3) |  |
| Adjusted hazard ratio for severe, critical or fatal COVID-19 (95% CI)<sup>‡</sup> | 0.20 (0.09 to 0.46) |  |
| Effectiveness of booster against severe, critical or fatal COVID-19 in % (95% CI)<sup>‡</sup> | 79.9 (54.2 to 91.2) |  |
| *Prior pre-omicron infection* |  |  |
| Adjusted hazard ratio for SARS-CoV-2 infection (95% CI)<sup>‡</sup> | 0.60 (0.56 to 0.64) |  |
| Effectiveness of booster against infection in % (95% CI)<sup>‡</sup> | 39.8 (35.7 to 43.6) |  |
| Adjusted hazard ratio for severe, critical or fatal COVID-19 (95% CI)<sup>‡</sup> | --<sup>§</sup> |  |
| Effectiveness of booster against severe, critical or fatal COVID-19 in % (95% CI)<sup>‡</sup> | --<sup>§</sup> |  |
| *Prior omicron infection* |  |  |
| Adjusted hazard ratio for SARS-CoV-2 infection (95% CI)<sup>‡</sup> | 0.77 (0.56 to 1.05) |  |
| Effectiveness of booster against infection in % (95% CI)<sup>‡</sup> | 23.2 (-4.5 to 43.7) |  |
| Adjusted hazard ratio for severe, critical or fatal COVID-19 (95% CI)<sup>‡</sup> | --<sup>§</sup> |  |
| Effectiveness of booster against severe, critical or fatal COVID-19 in % (95% CI)<sup>‡</sup> | --<sup>§</sup> |  |
| *Prior pre-omicron & omicron infections* |  |  |
| Adjusted hazard ratio for SARS-CoV-2 infection (95% CI)<sup>‡</sup> | 0.66 (0.09 to 4.94) |  |
| Effectiveness of booster against infection in % (95% CI)<sup>‡</sup> | 34.5 (-79.8 to 91.3) |  |
| Adjusted hazard ratio for severe, critical or fatal COVID-19 (95% CI)<sup>‡</sup> | --<sup>§</sup> |  |
| Effectiveness of booster against severe, critical or fatal COVID-19 in % (95% CI)<sup>‡</sup> | --<sup>§</sup> |  |
| **Estimates by clinical vulnerability to severe COVID-19** |  |  |
| *Persons less clinically vulnerable to severe COVID-19* |  |  |
| Adjusted hazard ratio for SARS-CoV-2 infection (95% CI)<sup>‡</sup> | 0.61 (0.60 to 0.63) |  |
| Effectiveness of booster against infection in % (95% CI)<sup>‡</sup> | 38.5 (37.3 to 39.8) |  |
| Adjusted hazard ratio for severe, critical or fatal COVID-19 (95% CI)<sup>‡</sup> | 0.47 (0.09 to 2.58)<sup>¶</sup> |  |
| Effectiveness of booster against severe, critical or fatal COVID-19 in % (95% CI)<sup>‡</sup> | 52.8 (-61.2 to 91.4)<sup>¶</sup> |  |
| *Persons more clinically vulnerable to severe COVID-19* |  |  |
| Adjusted hazard ratio for SARS-CoV-2 infection (95% CI)<sup>‡</sup> | 0.50 (0.48 to 0.52) |  |
| Effectiveness of booster against infection in % (95% CI)<sup>‡</sup> | 49.7 (47.8 to 51.6) |  |
| Adjusted hazard ratio for severe, critical or fatal COVID-19 (95% CI)<sup>‡</sup> | 0.16 (0.06 to 0.41) |  |
| Effectiveness of booster against severe, critical or fatal COVID-19 in % (95% CI)<sup>‡</sup> | 84.2 (58.8 to 93.9) |  |
| **Estimates by vaccine type** |  |  |
| *Vaccinated with BNT162b2* |  |  |
| Adjusted hazard ratio for SARS-CoV-2 infection (95% CI)<sup>‡</sup> | 0.56 (0.55 to 0.57) |  |
| Effectiveness of booster against infection in % (95% CI)<sup>‡</sup> | 44.0 (42.9 to 45.2) |  |
| Adjusted hazard ratio for severe, critical or fatal COVID-19 (95% CI)<sup>‡</sup> | 0.21 (0.09 to 0.47) |  |
| Effectiveness of booster against severe, critical or fatal COVID-19 in % (95% CI)<sup>‡</sup> | 79.4 (52.8 to 91.0) |  |
| *Vaccinated with mRNA-1273* |  |  |
| Adjusted hazard ratio for SARS-CoV-2 infection (95% CI)<sup>‡</sup> | 0.71 (0.68 to 0.74) |  |
| Effectiveness of booster against infection in % (95% CI)<sup>‡</sup> | 29.1 (26.4 to 31.6) |  |
| Adjusted hazard ratio for severe, critical or fatal COVID-19 (95% CI)<sup>‡</sup> | --<sup>§</sup> |  |
| Effectiveness of booster against severe, critical or fatal COVID-19 in % (95% CI)<sup>‡</sup> | --<sup>§</sup> |  |

The proportion of individuals who had a SARS-CoV-2 test during follow-up was 50.0% for the three-dose cohort and 45.3% for the two-dose cohort. The testing frequency was 1.13 and 0.98 tests per person, respectively. Adjusting the aHR in a sensitivity analysis additionally by the ratio of testing frequencies yielded a booster effectiveness against infection of 49.1% (95% CI: 48.2-50.0%).

### Booster effectiveness by prior infection status

Among persons with no prior infection, booster effectiveness was 41.2% (95% CI: 40.1-42.3%) against infection (Table 2, Table S2, and Figure 2A) and 79.9% (95% CI: 54.2-91.2%) against severe, critical, or fatal COVID-19 (Figure 2B).

**Figure 2:**
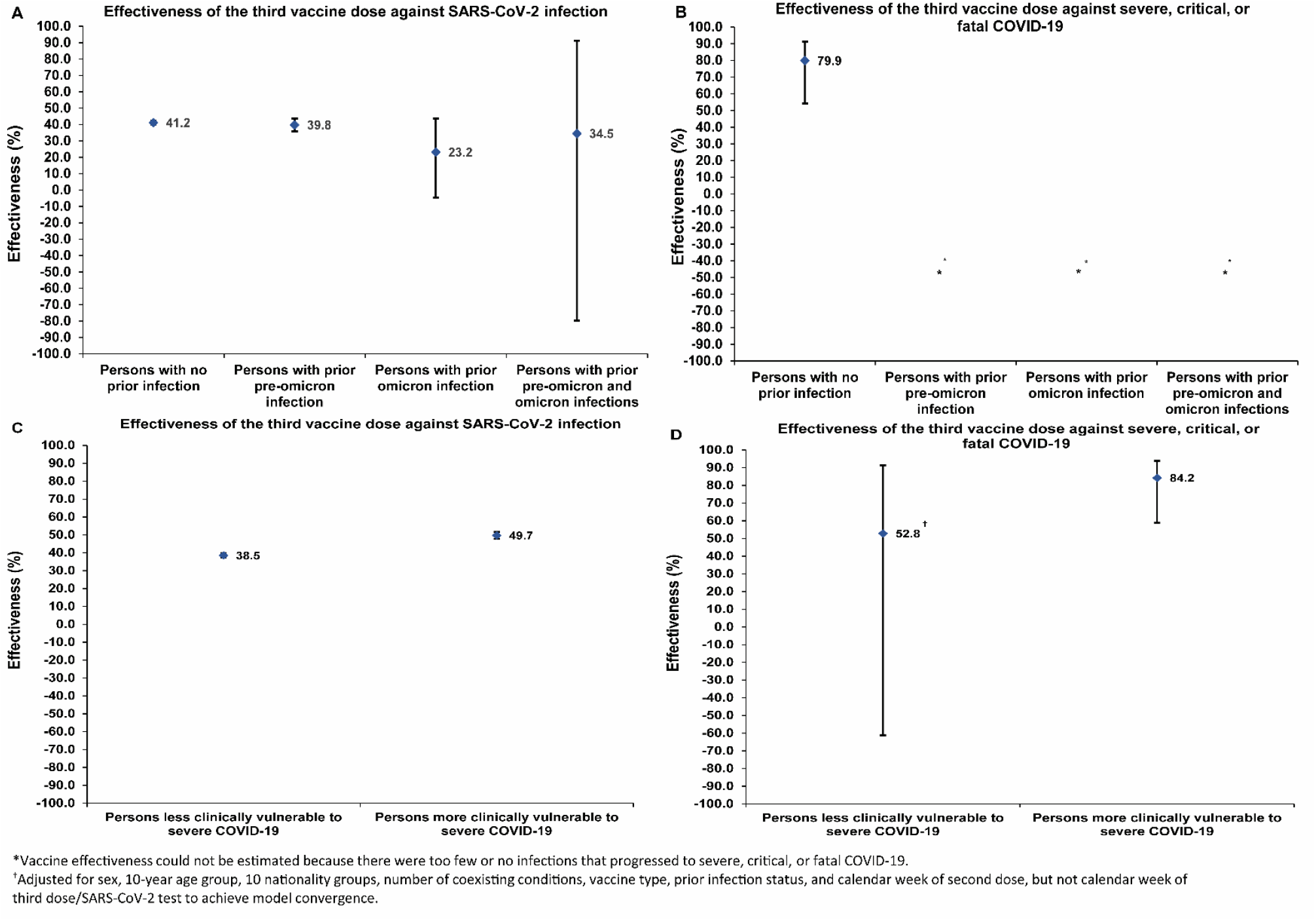
Booster effectiveness relative to primary series against SARS-CoV-2 infection and against severe, critical, or fatal COVID-19 by prior infection status (panels A and B, respectively) and by clinical vulnerability to severe COVID-19 (panels C and D, respectively).

Booster effectiveness against infection was 39.8% (95% CI: 35.7-43.6%) among persons with prior pre-omicron infection, 23.2% (95% CI: -4.5-43.7%) among persons with prior omicron infection, and 34.5% (95% CI: -79.8-91.3%) among persons with prior pre-omicron and omicron infections. Booster effectiveness against severe COVID-19 could not be estimated for each of these prior-infection subgroups because of too few severe COVID-19 cases.

### Booster effectiveness by clinical vulnerability to severe COVID-19

Among persons more clinically vulnerable to severe COVID-19, booster effectiveness was 49.7% (95% CI: 47.8-51.6%) against infection (Table 2, Table S2, and Figure 2C) and 84.2% (95% CI: 58.8-93.9%) against severe, critical, or fatal COVID-19 (Figure 2D).

Among persons less clinically vulnerable to severe COVID-19, booster effectiveness was 38.5% (95% CI: 37.3-39.8%) against infection and 52.8% (95% CI: -61.2-91.4%) against severe, critical, or fatal COVID-19. The wide 95% CI was a consequence of too few severe COVID-19 cases among persons less clinically vulnerable to severe COVID-19.

### Booster effectiveness by mRNA vaccine

Among persons vaccinated with BNT162b2, booster effectiveness was 44.0% (95% CI: 42.9-45.2%) against infection (Table 2, Table S2, and Figure S2A) and 79.4% (95% CI: 52.8-91.0%) against severe, critical, or fatal COVID-19 (Figure S2B).

Among persons vaccinated with mRNA-1273, booster effectiveness was 29.1% (95% CI: 26.4-31.6%) against infection. Effectiveness against severe, critical, or fatal COVID-19 could not be estimated because of too few severe cases.

### Waning of booster effectiveness

Booster effectiveness against infection was highest at 57.1% (95% CI: 55.9-58.3%) in the first month after the start of follow-up, but waned gradually thereafter and was modest at only 14.4% (95% CI: 7.3-20.9%) by the sixth month of follow-up (Figure 3). In the seventh month and thereafter, coincident with follow-up time during which BA.4/BA.5^32^ and BA.2.75*^33^ dominated incidence, effectiveness was progressively negative reaching -20.3% (95% CI: -55.0-29.0) after one year of follow-up.

**Figure 3:**
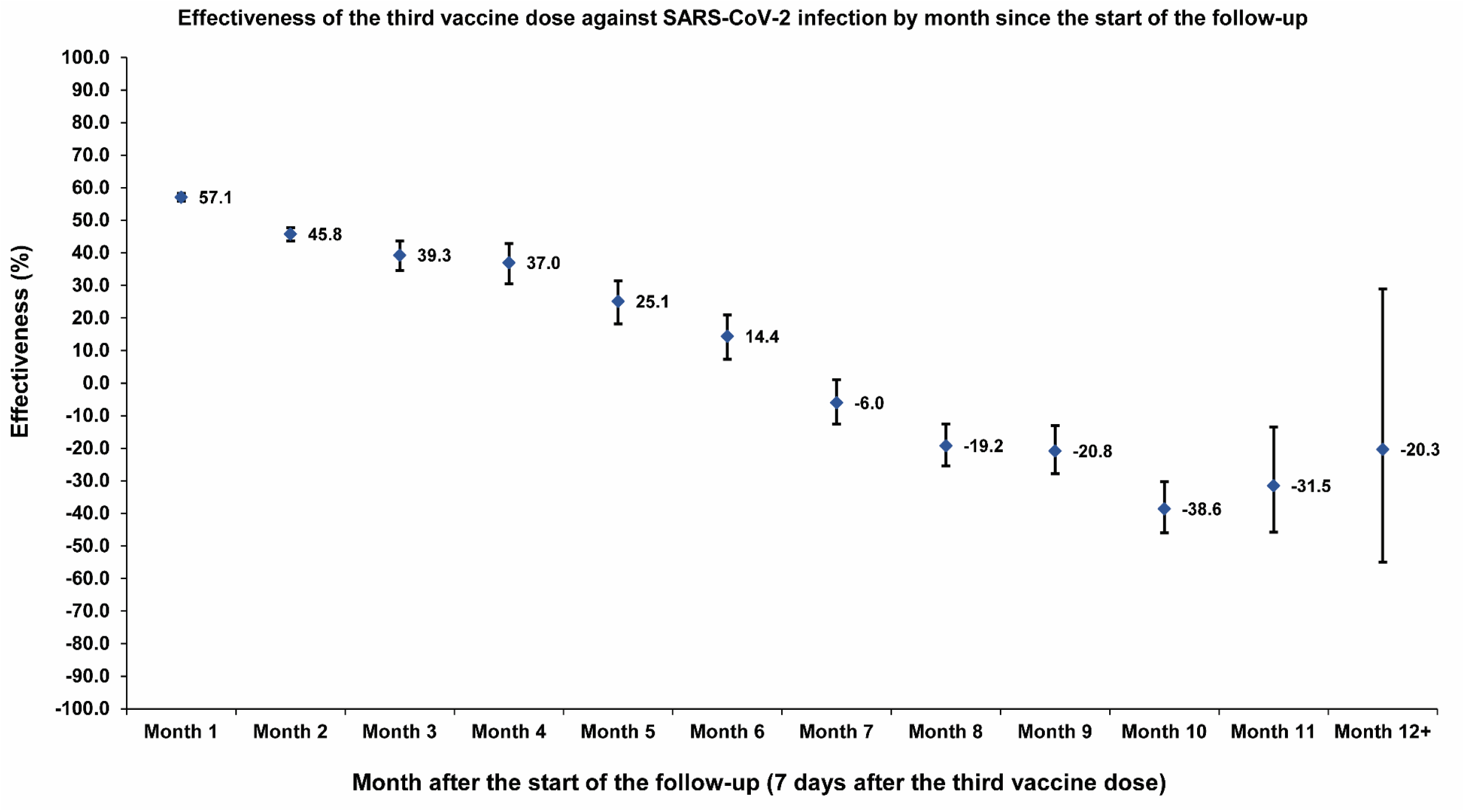
Booster effectiveness relative to primary series against SARS-CoV-2 infection by month since the start of the follow-up.

A similar pattern of waning of booster protection was observed irrespective of prior infection status (Figure 4A and 4B), clinical vulnerability to severe COVID-19 (Figure 4C and 4D), or vaccine type (Figure S3A and S3B). Effectiveness against severe, critical, or fatal COVID-19 could not be estimated by time interval of follow-up because of small number of severe COVID-19 cases.

**Figure 4:**
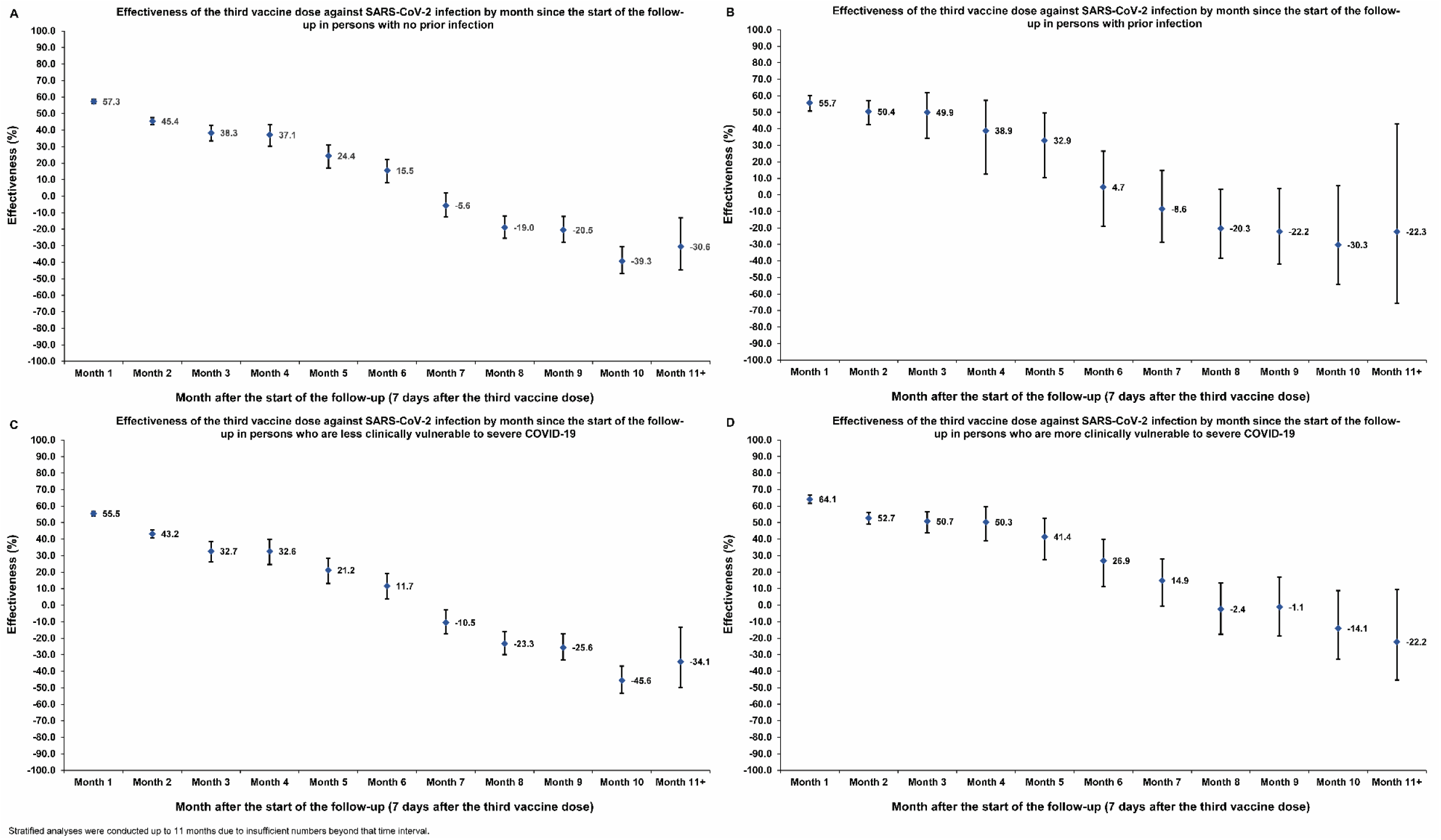
Booster effectiveness relative to primary series by month since the start of the follow-up against SARS-CoV-2 infection by prior infection status (panels A and B, respectively) and by clinical vulnerability to severe COVID-19 (panels C and D, respectively).

## Discussion

A third mRNA booster dose was associated with 41% reduction in incidence of infection and 81% reduction in incidence of severe COVID-19 over a year of follow-up. However, protection against infection waned gradually by month after the booster and was negligible by the sixth month. In the seventh month and thereafter, incidence of infection was higher among boosted persons compared to those with only the primary series, suggesting differential immune imprinting compromising protection in boosted persons against the newer omicron sublineages. There was no evidence for imprinting compromising protection against severe COVID-19, but the number of severe COVID-19 cases was too small to allow concrete estimation.

Evidence for imprinting was observed only after complete waning of booster effectiveness, and coincident with infections with new omicron subvariants BA.4/BA.5^32^ and BA.2.75*^33^ infections, consistent with a similar effect observed among cohorts who had a primary omicron infection, but different vaccination histories.^9^ The effect also seems consistent with emerging in vitro laboratory data.^7,34,35^ This suggests that imprinting affects a range of immune histories and is not restricted to specific ones. The effect was also observed irrespective of prior infection status, clinical vulnerability to severe COVID-19, or vaccine type, and was observed only after short-term booster protection had fully waned.

While imprinting is of concern when an antigenically divergent virus emerges, it likely does not negate the public health value of booster vaccinations. Imprinting affected protection against infection in the long-term, but the booster was protective against infection in the short-term. There was no evidence that imprinting affected protection against severe COVID-19 which remained high after over a year of follow-up. Imprinting was observed for boosters based on index-virus (pre-omicron) design,^36,37^ but it may not be observed for bivalent boosters, or may only be observed after a longer duration, as bivalent boosters may produce more effective antibodies for currently circulating viruses.

Ironically, imprinting may augment the need for repeated booster vaccination to blunt its effect. While imprinting has been observed for influenza,^38,39^ this has not undermined the public health value of seasonal influenza vaccinations,^40,41^ an outcome that may also apply for COVID-19 boosters. The findings, however, do accentuate the need for longer-term follow-up of boosted cohorts to understand the full extent of imprinting and its impact on both infection and severe disease.

The protective effects of boosters, relative to primary series, were similar irrespective of prior infection status highlighting the value of boosters even for those recovered from a prior infection, and irrespective of whether infections were due to pre-omicron or omicron viruses. The booster was associated with considerable protection against infection and high protection against severe COVID-19 among those more clinically vulnerable to severe COVID-19, underscoring the value of booster vaccinations for this population. While severe COVID-19 was rare among those less clinically vulnerable, the booster elicited considerable temporary protection against infection. Booster protection was higher among those more clinically vulnerable and those BNT162b2 vaccinated, but this may reflect slower waning of primary-series protection among those less clinically vulnerable and those mRNA-1273 vaccinated.^5,14,26^

This study has limitations. There were too few severe COVID-19 cases among some subgroups to allow estimation of effectiveness against severe COVID-19 in subgroup analyses or by time interval since the booster. Imprinting was observed among those with the longest time of follow-up, that is among those who first received the booster, but this population segment may not be representative of the wider population. Yet, imprinting was observed among those who received the booster at later months, and not only among those who were first eligible for the booster.

Testing frequency differed between cohorts, but sensitivity analysis adjusting for these differences showed similar findings. Home-based rapid-antigen testing is not documented (Section S1) and is not factored in these analyses. However, there is no reason to believe that home-based testing could have differentially affected the followed cohorts to alter study estimates. Matching was done while factoring key socio-demographic characteristics of the population,^15,20-23^ and this may also have controlled or reduced differences in home-based testing between cohorts.

As an observational study, investigated cohorts were neither blinded nor randomized, so unmeasured or uncontrolled confounding cannot be excluded. Although matching covered key factors affecting infection exposure,^15,20-23^ it was not possible for other factors such as geography or occupation, for which data were unavailable. However, Qatar is essentially a city state and infection incidence was broadly distributed across neighborhoods. Nearly 90% of Qatar’s population are expatriates from over 150 countries, who come here for employment.^15^ Nationality, age, and sex provide a powerful proxy for socio-economic status in this country.^15,20-23^ Nationality is strongly associated with occupation.^15,21-23^

The matching procedure used in this study was investigated in previous studies of different epidemiologic designs, and using control groups to test for null effects.^2,14,24-26^ These control groups have included unvaccinated cohorts versus vaccinated cohorts within two weeks of the first dose^2,24-26^ (when vaccine protection is negligible),^36,37^ and mRNA-1273-versus BNT162b2-vaccinated cohorts, also in the first two weeks after the first dose.^14^ These prior studies demonstrated at different times during the pandemic that this procedure provides adequate control of differences in infection exposure,^2,14,24-26^ suggesting that the matching strategy may also have controlled for differences in infection exposure in these studies. Analyses were implemented on Qatar’s total population and large samples, perhaps minimizing the likelihood of bias.

In conclusion, mRNA boosters reduced incidence of infection and severe COVID-19 over a year after vaccination, particularly among those clinically vulnerable to severe COVID-19. However, protection against infection waned after the booster, and eventually suggested an imprinting effect of compromised protection relative to the primary series. Both patterns of protection and imprinting were observed irrespective of prior infection status, clinical vulnerability to severe COVID-19, and vaccine type suggesting that the imprinting may affect a broad range of immune histories. While the imprinting is of concern, it may not negate the public health value of booster vaccination. There is need for longer-term follow-up of boosted cohorts to understand full extent of imprinting and its impact on infection and severe disease, particularly in the face of new emerging subvariants of SARS-CoV-2.

## Data Availability

The dataset of this study is a property of the Qatar Ministry of Public Health that was provided to the researchers through a restricted-access agreement that prevents sharing the dataset with a third party or publicly. Future access to this dataset can be considered through a direct application for data access to Her Excellency the Minister of Public Health (https://www.moph.gov.qa/english/Pages/default.aspx). Aggregate data are available within the manuscript and its Supplementary information.

## Author Contributons

HC, JSF, and LJA conceived and co-designed the study analyses. HC performed the statistical analyses and co-wrote the first draft of the article. LJA led the statistical analyses and co-wrote the first draft of the article. PT and MRH conducted multiplex, RT-qPCR variant screening and viral genome sequencing. PVC designed mass PCR testing to allow routine capture of SGTF variants and conducted viral genome sequencing. HY, HAK, and MS conducted viral genome sequencing. All authors contributed to data collection and acquisition, database development, discussion and interpretation of the results, and to the writing of the article. All authors have read and approved the final manuscript. Decision to publish the paper was by consensus among all authors.

## Acknowledgements

We acknowledge the many dedicated individuals at Hamad Medical Corporation, the Ministry of Public Health, the Primary Health Care Corporation, Qatar Biobank, Sidra Medicine, and Weill Cornell Medicine-Qatar for their diligent efforts and contributions to make this study possible. The authors are grateful for institutional salary support from the Biomedical Research Program and the Biostatistics, Epidemiology, and Biomathematics Research Core, both at Weill Cornell Medicine-Qatar, as well as for institutional salary support provided by the Ministry of Public Health, Hamad Medical Corporation, and Sidra Medicine. The authors are also grateful for the Qatar Genome Programme and Qatar University Biomedical Research Center for institutional support for the reagents needed for the viral genome sequencing. The funders of the study had no role in study design, data collection, data analysis, data interpretation, or writing of the article. Statements made herein are solely the responsibility of the authors.

## Supplementary Appendix

### Section S1: Further details on methods

#### Data sources and testing

Severe acute respiratory syndrome coronavirus 2 (SARS-CoV-2) testing in the healthcare system in Qatar is done at a mass scale, and mostly for routine reasons, where about 5% of the population are tested every week.^1,2^ About 75% of those diagnosed are diagnosed not because of appearance of symptoms, but because of routine testing.^1,2^ Every polymerase chain reaction (PCR) test and an increasing proportion of the facility-based rapid antigen tests conducted in Qatar, regardless of location or setting, are classified on the basis of symptoms and the reason for testing (clinical symptoms, contact tracing, surveys or random testing campaigns, individual requests, routine healthcare testing, pre-travel, at port of entry, or other). All facility-based testing done during follow-up in the present study was factored in the analyses of this study.

Rapid antigen test kits are available for purchase in pharmacies in Qatar, but outcome of home-based testing is not reported nor documented in the national databases. Since SARS-CoV-2-test outcomes are linked to specific public health measures, restrictions, and privileges, testing policy and guidelines stress facility-based testing as the core testing mechanism in the population.

While facility-based testing is provided free of charge or at low subsidized costs, depending on the reason for testing, home-based rapid antigen testing is de-emphasized and not supported as part of national policy. There is no reason to believe that home-based testing could have differentially affected the followed matched cohorts to affect our results.

The infection detection rate is defined as the cumulative number of documented infections, that is diagnosed and laboratory-confirmed infections, over the cumulative number of documented and undocumented infections. Serological surveys and other analyses suggest that a substantial proportion of infections in Qatar and elsewhere are undocumented.^3-9^ With absence of recent serological surveys in Qatar, it is difficult to estimate the current or recent infection detection rate, but mathematical modeling analyses and their recent updates suggest that at present no less than 50% of infections are never documented.^7,10^ However, there is no reason to believe that undocumented infections could have differentially affected the followed matched cohorts to affect our results.

Qatar has unusually young, diverse demographics, in that only 9% of its residents are ≥50 years of age, and 89% are expatriates from over 150 countries.^11,12^ Further descriptions of the study population and these national databases were reported previously.^1,2,12-14^

#### Comorbidity classification

Comorbidities were ascertained and classified based on the ICD-10 codes for chronic conditions as recorded in the electronic health record encounters of each individual in the Cerner-system national database that includes all citizens and residents registered in the national and universal public healthcare system. All encounters for each individual were analyzed to determine the comorbidity classification for that individual, as part of a recent national analysis to assess healthcare needs and resource allocation. The Cerner-system national database includes encounters starting from 2013, after this system was launched in Qatar. As long as each individual had at least one encounter with a specific comorbidity diagnosis since 2013, this person was classified with this comorbidity. Individuals who have comorbidities but never sought care in the public healthcare system, or seek care exclusively in private healthcare facilities, are classified as individuals with no comorbidity due to absence of recorded encounters for them.

### Section S2: Laboratory methods and variant ascertainment

#### Real-time reverse-transcription polymerase chain reaction testing

Nasopharyngeal and/or oropharyngeal swabs were collected for polymerase chain reaction (PCR) testing and placed in Universal Transport Medium (UTM). Aliquots of UTM were: 1) extracted on KingFisher Flex (Thermo Fisher Scientific, USA), MGISP-960 (MGI, China), or ExiPrep 96 Lite (Bioneer, South Korea) followed by testing with real-time reverse-transcription PCR (RT-qPCR) using TaqPath COVID-19 Combo Kits (Thermo Fisher Scientific, USA) on an ABI 7500 FAST (Thermo Fisher Scientific, USA); 2) tested directly on the Cepheid GeneXpert system using the Xpert Xpress SARS-CoV-2 (Cepheid, USA); or 3) loaded directly into a Roche cobas 6800 system and assayed with the cobas SARS-CoV-2 Test (Roche, Switzerland). The first assay targets the viral S, N, and ORF1ab gene regions. The second targets the viral N and E-gene regions, and the third targets the ORF1ab and E-gene regions.

All PCR testing was conducted at the Hamad Medical Corporation Central Laboratory or Sidra Medicine Laboratory, following standardized protocols.

#### Rapid antigen testing

Severe acute respiratory syndrome coronavirus 2 (SARS-CoV-2) antigen tests were performed on nasopharyngeal swabs using one of the following lateral flow antigen tests: Panbio COVID-19 Ag Rapid Test Device (Abbott, USA); SARS-CoV-2 Rapid Antigen Test (Roche, Switzerland); Standard Q COVID-19 Antigen Test (SD Biosensor, Korea); or CareStart COVID-19 Antigen Test (Access Bio, USA). All antigen tests were performed point-of-care according to each manufacturer’s instructions at public or private hospitals and clinics throughout Qatar with prior authorization and training by the Ministry of Public Health (MOPH). Antigen test results were electronically reported to the MOPH in real time using the Antigen Test Management System which is integrated with the national Coronavirus Disease 2019 (COVID-19) database.

#### Classification of infections by variant type

Surveillance for the severe acute respiratory syndrome coronavirus 2 (SARS-CoV-2) variants in Qatar is based on viral genome sequencing and multiplex real-time reverse-transcription polymerase chain reaction (RT-qPCR) variant screening^15^ of random positive clinical samples,^2,16-20^ complemented by deep sequencing of wastewater samples.^18,21,22^ Further details on the viral genome sequencing and multiplex RT-qPCR variant screening throughout the SARS-CoV-2 waves in Qatar can be found in previous publications.^1,2,14,16-20,23-28^

### Section S3: COVID-19 severity, criticality, and fatality classification

Classification of Coronavirus Disease 2019 (COVID-19) case severity (acute-care hospitalizations),^29^ criticality (intensive-care-unit hospitalizations),^29^ and fatality^30^ followed World Health Organization (WHO) guidelines. Assessments were made by trained medical personnel independent of study investigators and using individual chart reviews, as part of a national protocol applied to every hospitalized COVID-19 patient. Each hospitalized COVID-19 patient underwent an infection severity assessment every three days until discharge or death. We classified individuals who progressed to severe, critical, or fatal COVID-19 between the time of the documented infection and the end of the study based on their worst outcome, starting with death,^30^ followed by critical disease,^29^ and then severe disease.^29^

Severe COVID-19 disease was defined per WHO classification as a SARS-CoV-2 infected person with “oxygen saturation of <90% on room air, and/or respiratory rate of >30 breaths/minute in adults and children >5 years old (or ≥60 breaths/minute in children <2 months old or ≥50 breaths/minute in children 2-11 months old or ≥40 breaths/minute in children 1–5 years old), and/or signs of severe respiratory distress (accessory muscle use and inability to complete full sentences, and, in children, very severe chest wall indrawing, grunting, central cyanosis, or presence of any other general danger signs)”.^29^ Detailed WHO criteria for classifying Severe acute respiratory syndrome coronavirus 2 (SARS-CoV-2) infection severity can be found in the WHO technical report.^29^

Critical COVID-19 disease was defined per WHO classification as a SARS-CoV-2 infected person with “acute respiratory distress syndrome, sepsis, septic shock, or other conditions that would normally require the provision of life sustaining therapies such as mechanical ventilation (invasive or non-invasive) or vasopressor therapy”.^29^ Detailed WHO criteria for classifying SARS-CoV-2 infection criticality can be found in the WHO technical report.^29^

COVID-19 death was defined per WHO classification as “a death resulting from a clinically compatible illness, in a probable or confirmed COVID-19 case, unless there is a clear alternative cause of death that cannot be related to COVID-19 disease (e.g. trauma). There should be no period of complete recovery from COVID-19 between illness and death. A death due to COVID-19 may not be attributed to another disease (e.g. cancer) and should be counted independently of preexisting conditions that are suspected of triggering a severe course of COVID-19”. Detailed WHO criteria for classifying COVID-19 death can be found in the WHO technical report.^30^

**Table S1:** STROBE checklist for cohort studies.

|  | Item No | Recommendation | Main Text page |
| --- | --- | --- | --- |
| Title and abstract | 1 | (a) Indicate the study’s design with a commonly used term in the title or the abstract<br>(b) Provide in the abstract an informative and balanced summary of what was done and what was found | Abstract<br>Abstract |
| Introduction |  |  |  |
| Background/rationale | 2 | Explain the scientific background and rationale for the investigation being reported | Introduction |
| Objectives | 3 | State specific objectives, including any prespecified hypotheses | Introduction |
| Methods |  |  |  |
| Study design | 4 | Present key elements of study design early in the paper | Methods (‘Study design and cohorts’ & ‘Cohort matching and follow-up’) |
| Setting | 5 | Describe the setting, locations, and relevant dates, including periods of recruitment, exposure, follow-up, and data collection | Methods (‘Study design and cohorts’ & ‘Cohort matching and follow-up’), & Figure S1 & Sections S1-S3 in Supplementary Appendix |
| Participants | 6 | (a) Give the eligibility criteria, and the sources and methods of selection of participants. Describe methods of follow-up<br>(b) For matched studies, give matching criteria and number of exposed and unexposed | Methods (‘Study design and cohorts’ & ‘Cohort matching and follow-up’), & Figure S1 in Supplementary Appendix |
| Variables | 7 | Clearly define all outcomes, exposures, predictors, potential confounders, and effect modifiers. Give diagnostic criteria, if applicable | Methods (‘Study design and cohorts’ & ‘Cohort matching and follow-up’), Table 1, & Figure S1 & Sections S1-S3 in Supplementary Appendix. |
| Data sources/measurement | 8* | For each variable of interest, give sources of data and details of methods of assessment (measurement). Describe comparability of assessment methods if there is more than one group | Methods (‘Study population and data sources’ & ‘Statistical analysis’, paragraph 1), Table 1, & Sections S1-S3 in Supplementary Appendix |
| Bias | 9 | Describe any efforts to address potential sources of bias | Methods (‘Cohort matching and follow-up’) |
| Study size | 10 | Explain how the study size was arrived at | Figure S1 in Supplementary Appendix |
| Quantitative variables | 11 | Explain how quantitative variables were handled in the analyses. If applicable, describe which groupings were chosen and why | Methods (‘Cohort matching and follow-up’) & Table 1 |
| Statistical methods | 12 | (a) Describe all statistical methods, including those used to control for confounding | Methods (‘Study design and cohort’ & ‘Statistical analysis’) |
|  |  | (b) Describe any methods used to examine subgroups and interactions | Methods (‘Statistical analysis’) |
|  |  | (c) Explain how missing data were addressed | Not applicable, see Methods (‘Study population and data sources’) |
|  |  | (d) If applicable, explain how loss to follow-up was addressed | Not applicable, see Methods (‘Study population and data sources’) |
|  |  | (e) Describe any sensitivity analyses | Methods (‘Statistical analysis’, paragraph 2) |
| Results |  |  |  |
| Participants | 13* | (a) Report numbers of individuals at each stage of study—eg numbers potentially eligible, examined for eligibility, confirmed eligible, included in the study, completing follow-up, and analysed<br>(b) Give reasons for non-participation at each stage<br>(c) Consider use of a flow diagram | Figure S1 in Supplementary Appendix |
| Descriptive data | 14 | (a) Give characteristics of study participants (eg demographic, clinical, social) and information on exposures and potential confounders | Results (‘Overall booster effectiveness’, paragraphs 1-2), & Table 1 |
|  |  | (b) Indicate number of participants with missing data for each variable of interest | Not applicable, see Methods (‘Study population and data sources’) |
|  |  | (c) Summarise follow-up time (eg, average and total amount) | Results (‘Overall booster effectiveness’, paragraph 3), Figure 1, & Table 2 |
| Outcome data | 15 | Report numbers of outcome events or summary measures over time | Results (‘Overall booster effectiveness’, paragraph 4), Table 2, Figure 1, & Figure S1, in Supplementary Appendix. |
| Main results | 16 | (a) Give unadjusted estimates and, if applicable, confounder-adjusted estimates and their precision (eg, 95% confidence | Results (‘Overall booster effectiveness’, paragraphs 5-6, ‘Booster effectiveness by prior infection status’, ‘Booster |
|  |  | interval). Make clear which confounders were adjusted for and why they were included | effectiveness by clinical vulnerability to severe COVID-19', & 'Booster effectiveness by mRNA vaccine'), Table 2, Figure 2, & Table S2 & Figure S2 in Supplementary Appendix. |
|  |  | (b) Report category boundaries when continuous variables were categorized | Table 1 |
|  |  | (c) If relevant, consider translating estimates of relative risk into absolute risk for a meaningful time period | Not applicable |
| Other analyses | 17 | Report other analyses done—eg analyses of subgroups and interactions, and sensitivity analyses | Results ('Waning of booster effectiveness over time'), Figures 3 & 4, & Figure S3 in Supplementary Appendix. |
| Discussion |  |  |  |
| Key results | 18 | Summarise key results with reference to study objectives | Discussion, paragraphs 1-4 |
| Limitations | 19 | Discuss limitations of the study, taking into account sources of potential bias or imprecision. Discuss both direction and magnitude of any potential bias | Discussion, paragraphs 5-8 |
| Interpretation | 20 | Give a cautious overall interpretation of results considering objectives, limitations, multiplicity of analyses, results from similar studies, and other relevant evidence | Discussion, paragraph 9 |
| Generalisability | 21 | Discuss the generalisability (external validity) of the study results | Discussion, paragraphs 5-8 |
| Other information |  |  |  |
| Funding | 22 | Give the source of funding and the role of the funders for the present study and, if applicable, for the original study on which the present article is based | Acknowledgements |

**Figure S1:**
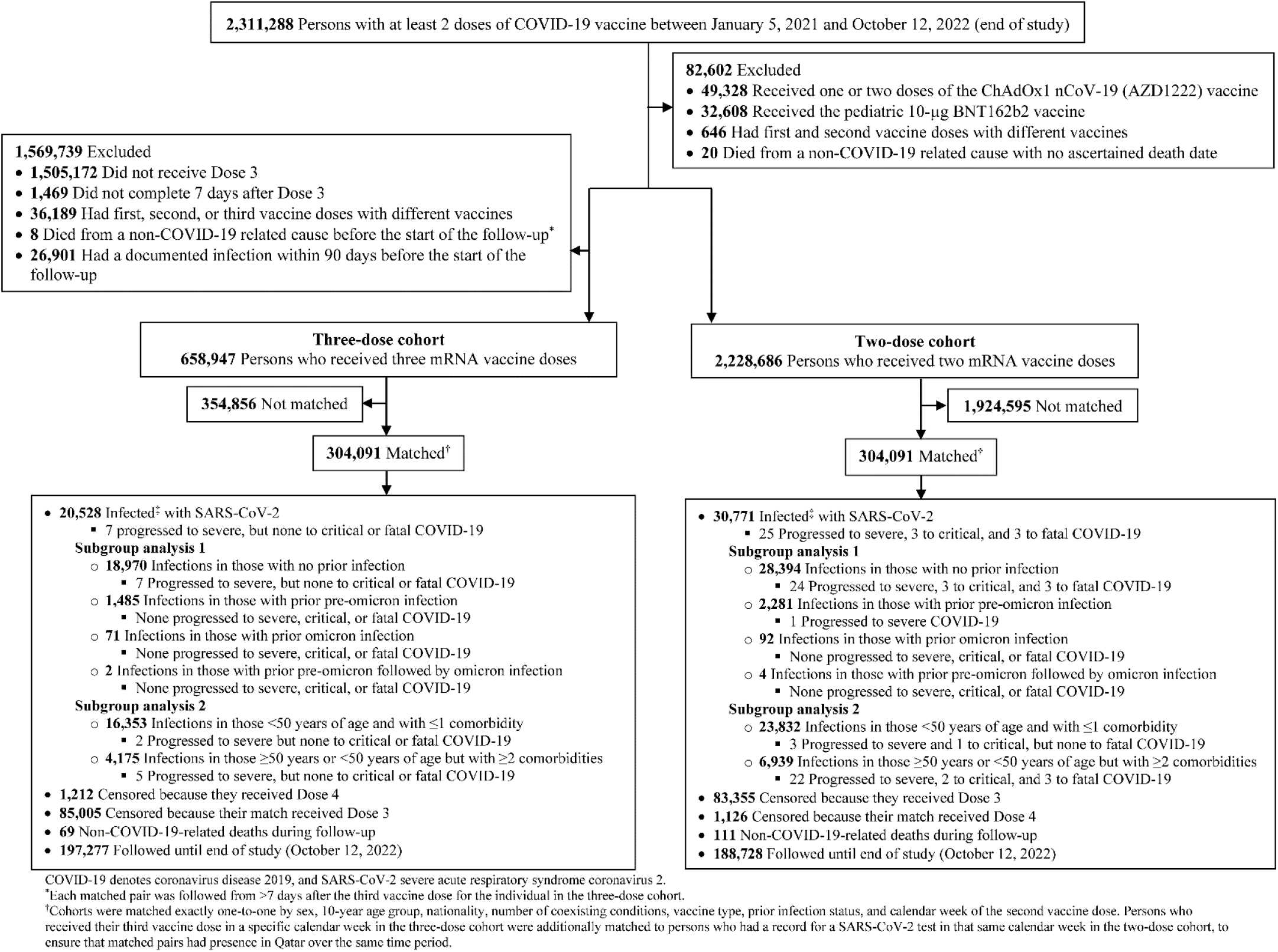
Cohort selection for investigating the effectiveness of mRNA booster (third dose) vaccination relative to that of primary-series (two-dose) vaccination.

**Table S2:**
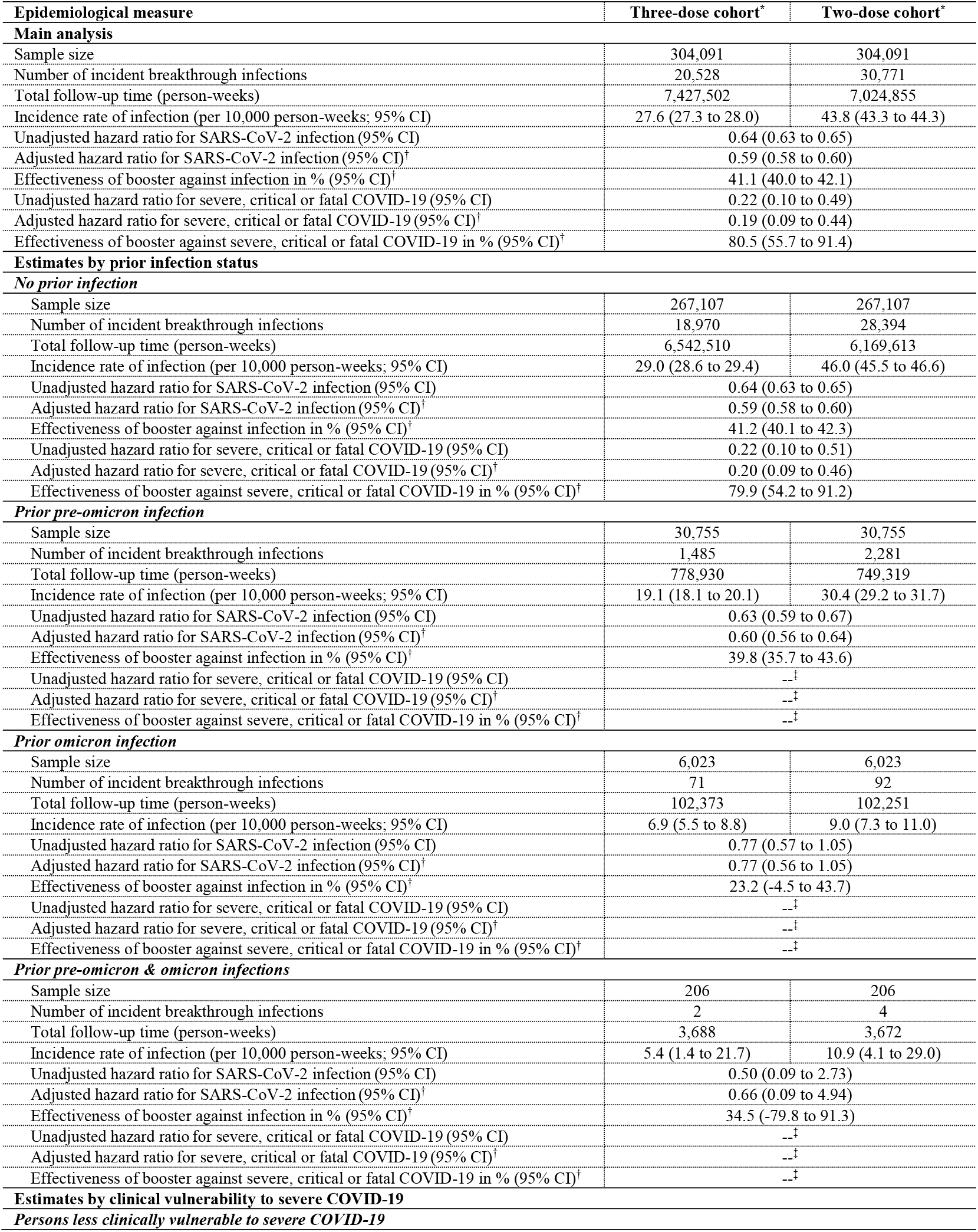
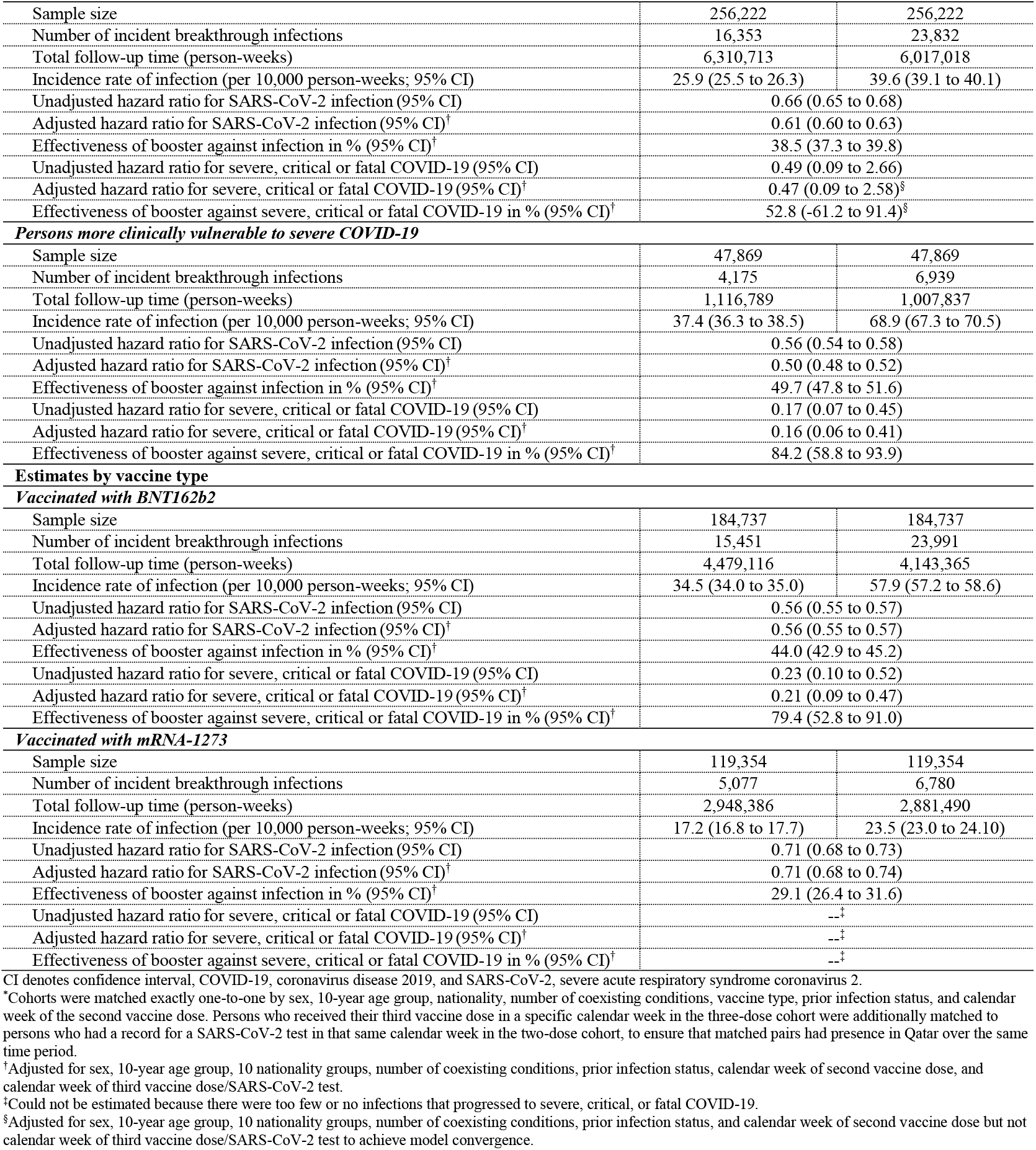
Detailed results for hazard ratios for incidence of SARS-CoV-2 infection and severe, critical or fatal COVID-19 in the three-dose cohort versus the two-dose cohort stratified by prior infection status, clinical vulnerability to severe COVID-19, and vaccine type.

| **Epidemiological measure** | **Three-dose cohort\*** | **Two-dose cohort\*** |
| --- | --- | --- |
| **Main analysis** |  |  |
| Sample size | 304,091 | 304,091 |
| Number of incident breakthrough infections | 20,528 | 30,771 |
| Total follow-up time (person-weeks) | 7,427,502 | 7,024,855 |
| Incidence rate of infection (per 10,000 person-weeks; 95% CI) | 27.6 (27.3 to 28.0) | 43.8 (43.3 to 44.3) |
| Unadjusted hazard ratio for SARS-CoV-2 infection (95% CI) | 0.64 (0.63 to 0.65) |  |
| Adjusted hazard ratio for SARS-CoV-2 infection (95% CI)<sup>†</sup> | 0.59 (0.58 to 0.60) |  |
| Effectiveness of booster against infection in % (95% CI)<sup>†</sup> | 41.1 (40.0 to 42.1) |  |
| Unadjusted hazard ratio for severe, critical or fatal COVID-19 (95% CI) | 0.22 (0.10 to 0.49) |  |
| Adjusted hazard ratio for severe, critical or fatal COVID-19 (95% CI)<sup>†</sup> | 0.19 (0.09 to 0.44) |  |
| Effectiveness of booster against severe, critical or fatal COVID-19 in % (95% CI)<sup>†</sup> | 80.5 (55.7 to 91.4) |  |
| **Estimates by prior infection status** |  |  |
| **No prior infection** |  |  |
| Sample size | 267,107 | 267,107 |
| Number of incident breakthrough infections | 18,970 | 28,394 |
| Total follow-up time (person-weeks) | 6,542,510 | 6,169,613 |
| Incidence rate of infection (per 10,000 person-weeks; 95% CI) | 29.0 (28.6 to 29.4) | 46.0 (45.5 to 46.6) |
| Unadjusted hazard ratio for SARS-CoV-2 infection (95% CI) | 0.64 (0.63 to 0.65) |  |
| Adjusted hazard ratio for SARS-CoV-2 infection (95% CI)<sup>†</sup> | 0.59 (0.58 to 0.60) |  |
| Effectiveness of booster against infection in % (95% CI)<sup>†</sup> | 41.2 (40.1 to 42.3) |  |
| Unadjusted hazard ratio for severe, critical or fatal COVID-19 (95% CI) | 0.22 (0.10 to 0.51) |  |
| Adjusted hazard ratio for severe, critical or fatal COVID-19 (95% CI)<sup>†</sup> | 0.20 (0.09 to 0.46) |  |
| Effectiveness of booster against severe, critical or fatal COVID-19 in % (95% CI)<sup>†</sup> | 79.9 (54.2 to 91.2) |  |
| **Prior pre-omicron infection** |  |  |
| Sample size | 30,755 | 30,755 |
| Number of incident breakthrough infections | 1,485 | 2,281 |
| Total follow-up time (person-weeks) | 778,930 | 749,319 |
| Incidence rate of infection (per 10,000 person-weeks; 95% CI) | 19.1 (18.1 to 20.1) | 30.4 (29.2 to 31.7) |
| Unadjusted hazard ratio for SARS-CoV-2 infection (95% CI) | 0.63 (0.59 to 0.67) |  |
| Adjusted hazard ratio for SARS-CoV-2 infection (95% CI)<sup>†</sup> | 0.60 (0.56 to 0.64) |  |
| Effectiveness of booster against infection in % (95% CI)<sup>†</sup> | 39.8 (35.7 to 43.6) |  |
| Unadjusted hazard ratio for severe, critical or fatal COVID-19 (95% CI) | --<sup>‡</sup> |  |
| Adjusted hazard ratio for severe, critical or fatal COVID-19 (95% CI)<sup>†</sup> | --<sup>‡</sup> |  |
| Effectiveness of booster against severe, critical or fatal COVID-19 in % (95% CI)<sup>†</sup> | --<sup>‡</sup> |  |
| **Prior omicron infection** |  |  |
| Sample size | 6,023 | 6,023 |
| Number of incident breakthrough infections | 71 | 92 |
| Total follow-up time (person-weeks) | 102,373 | 102,251 |
| Incidence rate of infection (per 10,000 person-weeks; 95% CI) | 6.9 (5.5 to 8.8) | 9.0 (7.3 to 11.0) |
| Unadjusted hazard ratio for SARS-CoV-2 infection (95% CI) | 0.77 (0.57 to 1.05) |  |
| Adjusted hazard ratio for SARS-CoV-2 infection (95% CI)<sup>†</sup> | 0.77 (0.56 to 1.05) |  |
| Effectiveness of booster against infection in % (95% CI)<sup>†</sup> | 23.2 (-4.5 to 43.7) |  |
| Unadjusted hazard ratio for severe, critical or fatal COVID-19 (95% CI) | --<sup>‡</sup> |  |
| Adjusted hazard ratio for severe, critical or fatal COVID-19 (95% CI)<sup>†</sup> | --<sup>‡</sup> |  |
| Effectiveness of booster against severe, critical or fatal COVID-19 in % (95% CI)<sup>†</sup> | --<sup>‡</sup> |  |
| **Prior pre-omicron & omicron infections** |  |  |
| Sample size | 206 | 206 |
| Number of incident breakthrough infections | 2 | 4 |
| Total follow-up time (person-weeks) | 3,688 | 3,672 |
| Incidence rate of infection (per 10,000 person-weeks; 95% CI) | 5.4 (1.4 to 21.7) | 10.9 (4.1 to 29.0) |
| Unadjusted hazard ratio for SARS-CoV-2 infection (95% CI) | 0.50 (0.09 to 2.73) |  |
| Adjusted hazard ratio for SARS-CoV-2 infection (95% CI)<sup>†</sup> | 0.66 (0.09 to 4.94) |  |
| Effectiveness of booster against infection in % (95% CI)<sup>†</sup> | 34.5 (-79.8 to 91.3) |  |
| Unadjusted hazard ratio for severe, critical or fatal COVID-19 (95% CI) | --<sup>‡</sup> |  |
| Adjusted hazard ratio for severe, critical or fatal COVID-19 (95% CI)<sup>†</sup> | --<sup>‡</sup> |  |
| Effectiveness of booster against severe, critical or fatal COVID-19 in % (95% CI)<sup>†</sup> | --<sup>‡</sup> |  |
| **Estimates by clinical vulnerability to severe COVID-19** |  |  |
| **Persons less clinically vulnerable to severe COVID-19** |  |  |
| Sample size | 256,222 | 256,222 |
| Number of incident breakthrough infections | 16,353 | 23,832 |
| Total follow-up time (person-weeks) | 6,310,713 | 6,017,018 |
| Incidence rate of infection (per 10,000 person-weeks; 95% CI) | 25.9 (25.5 to 26.3) | 39.6 (39.1 to 40.1) |
| Unadjusted hazard ratio for SARS-CoV-2 infection (95% CI) | 0.66 (0.65 to 0.68) |  |
| Adjusted hazard ratio for SARS-CoV-2 infection (95% CI)<sup>†</sup> | 0.61 (0.60 to 0.63) |  |
| Effectiveness of booster against infection in % (95% CI)<sup>†</sup> | 38.5 (37.3 to 39.8) |  |
| Unadjusted hazard ratio for severe, critical or fatal COVID-19 (95% CI) | 0.49 (0.09 to 2.66) |  |
| Adjusted hazard ratio for severe, critical or fatal COVID-19 (95% CI)<sup>†</sup> | 0.47 (0.09 to 2.58)<sup>§</sup> |  |
| Effectiveness of booster against severe, critical or fatal COVID-19 in % (95% CI)<sup>†</sup> | 52.8 (-61.2 to 91.4)<sup>§</sup> |  |
| ***Persons more clinically vulnerable to severe COVID-19*** |  |  |
| Sample size | 47,869 | 47,869 |
| Number of incident breakthrough infections | 4,175 | 6,939 |
| Total follow-up time (person-weeks) | 1,116,789 | 1,007,837 |
| Incidence rate of infection (per 10,000 person-weeks; 95% CI) | 37.4 (36.3 to 38.5) | 68.9 (67.3 to 70.5) |
| Unadjusted hazard ratio for SARS-CoV-2 infection (95% CI) | 0.56 (0.54 to 0.58) |  |
| Adjusted hazard ratio for SARS-CoV-2 infection (95% CI)<sup>†</sup> | 0.50 (0.48 to 0.52) |  |
| Effectiveness of booster against infection in % (95% CI)<sup>†</sup> | 49.7 (47.8 to 51.6) |  |
| Unadjusted hazard ratio for severe, critical or fatal COVID-19 (95% CI) | 0.17 (0.07 to 0.45) |  |
| Adjusted hazard ratio for severe, critical or fatal COVID-19 (95% CI)<sup>†</sup> | 0.16 (0.06 to 0.41) |  |
| Effectiveness of booster against severe, critical or fatal COVID-19 in % (95% CI)<sup>†</sup> | 84.2 (58.8 to 93.9) |  |
| **Estimates by vaccine type** |  |  |
| ***Vaccinated with BNT162b2*** |  |  |
| Sample size | 184,737 | 184,737 |
| Number of incident breakthrough infections | 15,451 | 23,991 |
| Total follow-up time (person-weeks) | 4,479,116 | 4,143,365 |
| Incidence rate of infection (per 10,000 person-weeks; 95% CI) | 34.5 (34.0 to 35.0) | 57.9 (57.2 to 58.6) |
| Unadjusted hazard ratio for SARS-CoV-2 infection (95% CI) | 0.56 (0.55 to 0.57) |  |
| Adjusted hazard ratio for SARS-CoV-2 infection (95% CI)<sup>†</sup> | 0.56 (0.55 to 0.57) |  |
| Effectiveness of booster against infection in % (95% CI)<sup>†</sup> | 44.0 (42.9 to 45.2) |  |
| Unadjusted hazard ratio for severe, critical or fatal COVID-19 (95% CI) | 0.23 (0.10 to 0.52) |  |
| Adjusted hazard ratio for severe, critical or fatal COVID-19 (95% CI)<sup>†</sup> | 0.21 (0.09 to 0.47) |  |
| Effectiveness of booster against severe, critical or fatal COVID-19 in % (95% CI)<sup>†</sup> | 79.4 (52.8 to 91.0) |  |
| ***Vaccinated with mRNA-1273*** |  |  |
| Sample size | 119,354 | 119,354 |
| Number of incident breakthrough infections | 5,077 | 6,780 |
| Total follow-up time (person-weeks) | 2,948,386 | 2,881,490 |
| Incidence rate of infection (per 10,000 person-weeks; 95% CI) | 17.2 (16.8 to 17.7) | 23.5 (23.0 to 24.10) |
| Unadjusted hazard ratio for SARS-CoV-2 infection (95% CI) | 0.71 (0.68 to 0.73) |  |
| Adjusted hazard ratio for SARS-CoV-2 infection (95% CI)<sup>†</sup> | 0.71 (0.68 to 0.74) |  |
| Effectiveness of booster against infection in % (95% CI)<sup>†</sup> | 29.1 (26.4 to 31.6) |  |
| Unadjusted hazard ratio for severe, critical or fatal COVID-19 (95% CI) | --<sup>‡</sup> |  |
| Adjusted hazard ratio for severe, critical or fatal COVID-19 (95% CI)<sup>†</sup> | --<sup>‡</sup> |  |
| Effectiveness of booster against severe, critical or fatal COVID-19 in % (95% CI)<sup>†</sup> | --<sup>‡</sup> |  |

**Figure S2:**
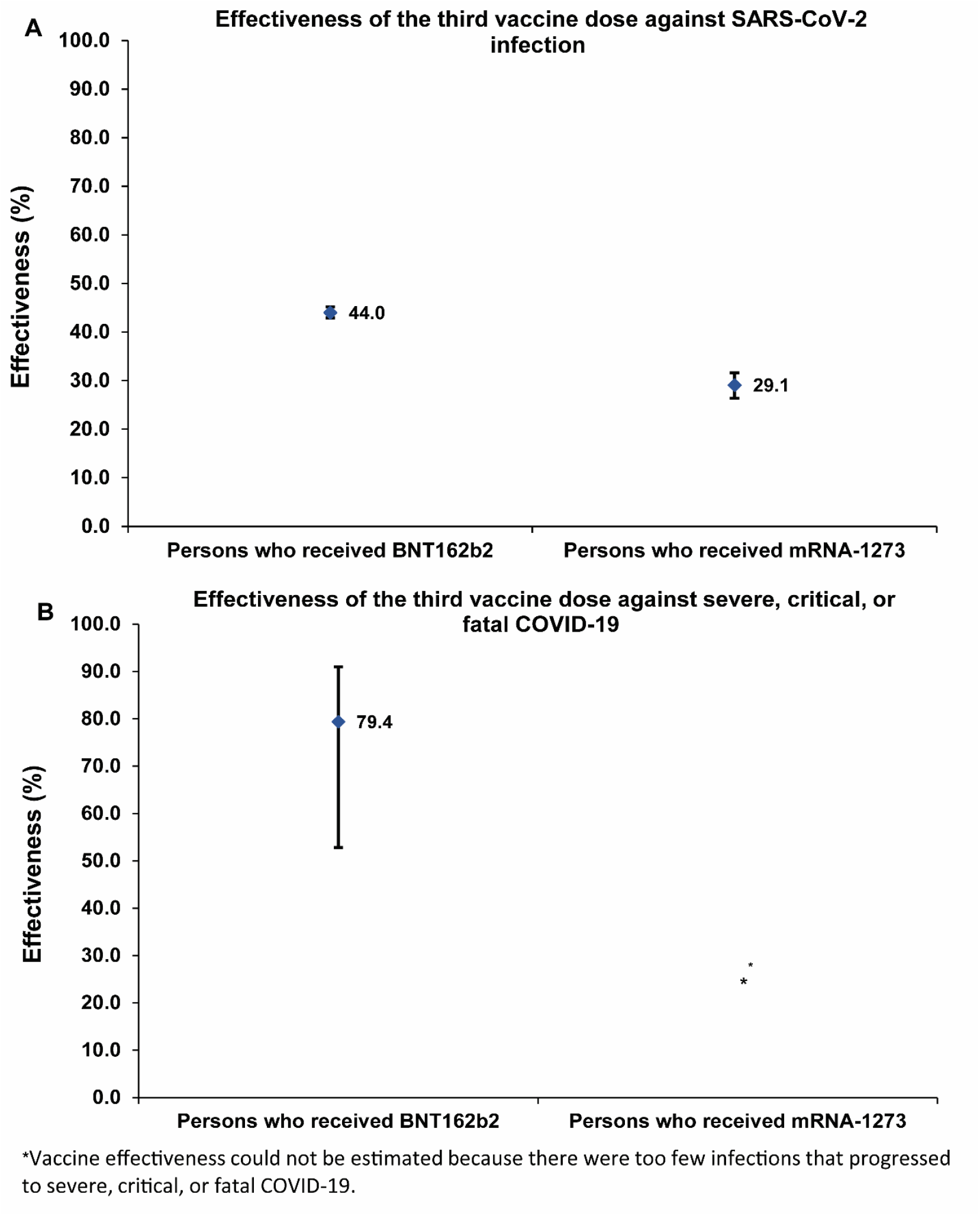
Booster effectiveness relative to primary series by mRNA vaccine type A) against SARS-CoV-2 infection and B) against severe, critical, or fatal COVID-19.

**Figure S3:**
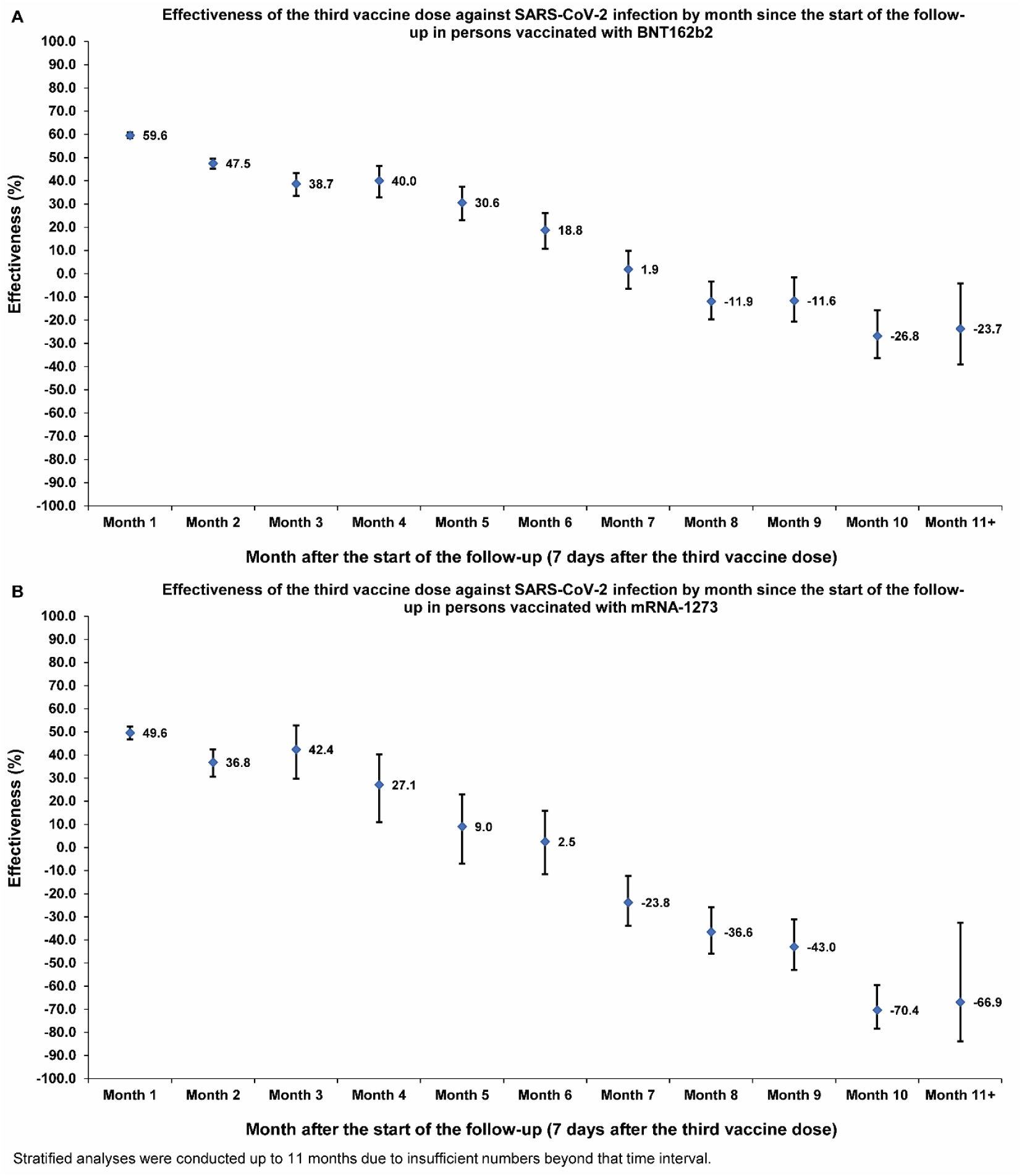
Booster effectiveness relative to primary series against SARS-CoV-2 infection by month since the start of the follow-up for each of BNT162b2 (A) and mRNA-1273 (B) vaccines.

## References

1. Abu-Raddad LJ, Chemaitelly H, Bertollini R, National Study Group for C-V. Waning mRNA-1273 Vaccine Effectiveness against SARS-CoV-2 Infection in Qatar. N Engl J Med 2022;386:1091–3.

2. Chemaitelly H, Tang P, Hasan MR, et al. Waning of BNT162b2 Vaccine Protection against SARS-CoV-2 Infection in Qatar. N Engl J Med 2021;385:e83.

3. Chemaitelly H, Nagelkerke N, Ayoub HH, et al. Duration of immune protection of SARS-CoV-2 natural infection against reinfection. J Travel Med 2022.

4. Tartof SY, Slezak JM, Puzniak L, et al. Effectiveness and durability of BNT162b2 vaccine against hospital and emergency department admissions due to SARS-CoV-2 omicron sub-lineages BA.1 and BA.2 in a large health system in the USA: a test-negative, case-control study. Lancet Respir Med 2022.

5. Abu-Raddad LJ, Chemaitelly H, Ayoub HH, et al. Effect of mRNA Vaccine Boosters against SARS-CoV-2 Omicron Infection in Qatar. N Engl J Med 2022;386:1804–16.

6. Chemaitelly H, Ayoub HH, AlMukdad S, et al. Duration of mRNA vaccine protection against SARS-CoV-2 Omicron BA.1 and BA.2 subvariants in Qatar. Nat Commun 2022;13:3082.

7. Reynolds CJ, Pade C, Gibbons JM, et al. Immune boosting by B.1.1.529 (Omicron) depends on previous SARS-CoV-2 exposure. Science 2022:eabq1841.

8. Chemaitelly H, Ayoub HH, Tang P, et al. Immune Imprinting and Protection against Repeat Reinfection with SARS-CoV-2. N Engl J Med 2022.

9. Chemaitelly H, Ayoub HH, Tang P, et al. COVID-19 primary series and booster vaccination and immune imprinting. medRxiv 2022:2022.10.31.22281756.

10. Röltgen K, Nielsen SCA, Silva O, et al. Immune imprinting, breadth of variant recognition, and germinal center response in human SARS-CoV-2 infection and vaccination. Cell 2022;185:1025-40.e14.

11. World Health Organization. COVID-19 clinical management: living guidance. Available from: https://www.who.int/publications/i/item/WHO-2019-nCoV-clinical-2021-1. Accessed on: May 15, 2021. 2021.

12. World Health Organization. International guidelines for certification and classification (coding) of COVID-19 as cause of death. Available from: https://www.who.int/classifications/icd/Guidelines_Cause_of_Death_COVID-19-20200420-EN.pdf?ua=1. xDocument Number: WHO/HQ/DDI/DNA/CAT. Accessed on May 15, 2021. 2020.

13. Altarawneh HN, Chemaitelly H, Ayoub HH, et al. Effects of Previous Infection and Vaccination on Symptomatic Omicron Infections. N Engl J Med 2022;387:21–34.

14. Abu-Raddad LJ, Chemaitelly H, Bertollini R, National Study Group for Covid Vaccination. Effectiveness of mRNA-1273 and BNT162b2 Vaccines in Qatar. N Engl J Med 2022;386:799–800.

15. Abu-Raddad LJ, Chemaitelly H, Ayoub HH, et al. Characterizing the Qatar advanced-phase SARS-CoV-2 epidemic. Sci Rep 2021;11:6233.

16. Chemaitelly H, Bertollini R, Abu-Raddad LJ, National Study Group for Covid Epidemiology. Efficacy of Natural Immunity against SARS-CoV-2 Reinfection with the Beta Variant. N Engl J Med 2021;385:2585–6.

17. Hernan MA, Robins JM. Using Big Data to Emulate a Target Trial When a Randomized Trial Is Not Available. Am J Epidemiol 2016;183:758–64.

18. Altarawneh HN, Chemaitelly H, Hasan MR, et al. Protection against the Omicron Variant from Previous SARS-CoV-2 Infection. N Engl J Med 2022;386:1288–90.

19. Seedat S, Chemaitelly H, Ayoub HH, et al. SARS-CoV-2 infection hospitalization, severity, criticality, and fatality rates in Qatar. Sci Rep 2021;11:18182.

20. Ayoub HH, Chemaitelly H, Seedat S, et al. Mathematical modeling of the SARS-CoV-2 epidemic in Qatar and its impact on the national response to COVID-19. J Glob Health 2021;11:05005.

21. Coyle PV, Chemaitelly H, Ben Hadj Kacem MA, et al. SARS-CoV-2 seroprevalence in the urban population of Qatar: An analysis of antibody testing on a sample of 112,941 individuals. iScience 2021;24:102646.

22. Al-Thani MH, Farag E, Bertollini R, et al. SARS-CoV-2 Infection Is at Herd Immunity in the Majority Segment of the Population of Qatar. Open Forum Infect Dis 2021;8:ofab221.

23. Jeremijenko A, Chemaitelly H, Ayoub HH, et al. Herd Immunity against Severe Acute Respiratory Syndrome Coronavirus 2 Infection in 10 Communities, Qatar. Emerg Infect Dis 2021;27:1343–52.

24. Abu-Raddad LJ, Chemaitelly H, Yassine HM, et al. Pfizer-BioNTech mRNA BNT162b2 Covid-19 vaccine protection against variants of concern after one versus two doses. J Travel Med 2021;28.

25. Chemaitelly H, Yassine HM, Benslimane FM, et al. mRNA-1273 COVID-19 vaccine effectiveness against the B.1.1.7 and B.1.351 variants and severe COVID-19 disease in Qatar. Nat Med 2021;27:1614–21.

26. Abu-Raddad LJ, Chemaitelly H, Bertollini R, National Study Group for Covid Vaccination. Waning mRNA-1273 Vaccine Effectiveness against SARS-CoV-2 Infection in Qatar. N Engl J Med 2022;386:1091–3.

27. Kojima N, Shrestha NK, Klausner JD. A Systematic Review of the Protective Effect of Prior SARS-CoV-2 Infection on Repeat Infection. Eval Health Prof 2021;44:327–32.

28. Pilz S, Theiler-Schwetz V, Trummer C, Krause R, Ioannidis JPA. SARS-CoV-2 reinfections: Overview of efficacy and duration of natural and hybrid immunity. Environ Res 2022:112911.

29. Barda N, Dagan N, Cohen C, et al. Effectiveness of a third dose of the BNT162b2 mRNA COVID-19 vaccine for preventing severe outcomes in Israel: an observational study. Lancet 2021;398:2093–100.

30. Austin PC. Using the Standardized Difference to Compare the Prevalence of a Binary Variable Between Two Groups in Observational Research. Communications in Statistics - Simulation and Computation 2009;38:1228–34.

31. Chemaitelly H, Ayoub HH, Coyle P, et al. Protection of Omicron sub-lineage infection against reinfection with another Omicron sub-lineage. Nat Commun 2022;13:4675.

32. Altarawneh HN, Chemaitelly H, Ayoub HH, et al. Protective Effect of Previous SARS-CoV-2 Infection against Omicron BA.4 and BA.5 Subvariants. N Engl J Med 2022.

33. Chemaitelly H, Tang P, Coyle P, et al. Protection against reinfection with SARS-CoV-2 omicron BA.2.75<sup>*</sup> sublineage. medRxiv 2022:2022.10.29.22281606.

34. Collier A-r, Miller J, Hachmann N, et al. Immunogenicity of the BA.5 Bivalent mRNA Vaccine Boosters. bioRxiv 2022:2022.10.24.513619.

35. Wang Q, Bowen A, Valdez R, et al. Antibody responses to Omicron BA.4/BA.5 bivalent mRNA vaccine booster shot. bioRxiv 2022:2022.10.22.513349.

36. Polack FP, Thomas SJ, Kitchin N, et al. Safety and Efficacy of the BNT162b2 mRNA Covid-19 Vaccine. N Engl J Med 2020;383:2603–15.

37. Baden LR, El Sahly HM, Essink B, et al. Efficacy and Safety of the mRNA-1273 SARS-CoV-2 Vaccine. N Engl J Med 2021;384:403–16.

38. Janjua NZ, Skowronski DM, Hottes TS, et al. Seasonal influenza vaccine and increased risk of pandemic A/H1N1-related illness: first detection of the association in British Columbia, Canada. Clin Infect Dis 2010;51:1017–27.

39. Skowronski DM, Sabaiduc S, Leir S, et al. Paradoxical clade- and age-specific vaccine effectiveness during the 2018/19 influenza A(H3N2) epidemic in Canada: potential imprint-regulated effect of vaccine (I-REV). Euro Surveill 2019;24.

40. Behrouzi B, Bhatt DL, Cannon CP, et al. Association of Influenza Vaccination With Cardiovascular Risk: A Meta-analysis. JAMA network open 2022;5:e228873.

41. Chung H, Buchan SA, Campigotto A, et al. Influenza Vaccine Effectiveness Against All-Cause Mortality Following Laboratory-Confirmed Influenza in Older Adults, 2010–2011 to 2015–2016 Seasons in Ontario, Canada. Clinical Infectious Diseases 2020;73:e1191–e9.

## References

1. Altarawneh HN, Chemaitelly H, Ayoub HH, et al. Effects of Previous Infection and Vaccination on Symptomatic Omicron Infections. N Engl J Med 2022;387:21–34.

3. Al-Thani MH, Farag E, Bertollini R, et al. SARS-CoV-2 infection is at herd immunity in the majority segment of the population of Qatar. Open Forum Infectious Diseases 2021.

4. Coyle PV, Chemaitelly H, Ben Hadj Kacem MA, et al. SARS-CoV-2 seroprevalence in the urban population of Qatar: An analysis of antibody testing on a sample of 112,941 individuals. iScience 2021;24:102646.

5. Jeremijenko A, Chemaitelly H, Ayoub HH, et al. Herd Immunity against Severe Acute Respiratory Syndrome Coronavirus 2 Infection in 10 Communities, Qatar. Emerg Infect Dis 2021;27:1343–52.

6. Seedat S, Chemaitelly H, Ayoub HH, et al. SARS-CoV-2 infection hospitalization, severity, criticality, and fatality rates in Qatar. Sci Rep 2021;11:18182.

7. Ayoub HH, Chemaitelly H, Seedat S, et al. Mathematical modeling of the SARS-CoV-2 epidemic in Qatar and its impact on the national response to COVID-19. J Glob Health 2021;11:05005.

8. Ayoub HH, Mumtaz GR, Seedat S, Makhoul M, Chemaitelly H, Abu-Raddad LJ. Estimates of global SARS-CoV-2 infection exposure, infection morbidity, and infection mortality rates in 2020. Glob Epidemiol 2021;3:100068.

9. Angulo FJ, Finelli L, Swerdlow DL. Estimation of US SARS-CoV-2 Infections, Symptomatic Infections, Hospitalizations, and Deaths Using Seroprevalence Surveys. JAMA Netw Open 2021;4:e2033706.

10. Ayoub HH, Chemaitelly H, Makhoul M, et al. Epidemiological impact of prioritising SARS-CoV-2 vaccination by antibody status: mathematical modelling analyses. BMJ Innov 2021;7:327–36.

11. Planning and Statistics Authority-State of Qatar. Qatar Monthly Statistics. Available from: https://www.psa.gov.qa/en/pages/default.aspx. Accessed on: May 26, 2020. 2020.

12. Abu-Raddad LJ, Chemaitelly H, Ayoub HH, et al. Characterizing the Qatar advanced-phase SARS-CoV-2 epidemic. Sci Rep 2021;11:6233.

13. Chemaitelly H, Bertollini R, Abu-Raddad LJ, National Study Group for Covid Epidemiology. Efficacy of Natural Immunity against SARS-CoV-2 Reinfection with the Beta Variant. N Engl J Med 2021;385:2585–6.

14. Abu-Raddad LJ, Chemaitelly H, Ayoub HH, et al. Effect of mRNA Vaccine Boosters against SARS-CoV-2 Omicron Infection in Qatar. N Engl J Med 2022;386:1804–16.

15. Multiplexed RT-qPCR to screen for SARS-COV-2 B.1.1.7, B.1.351, and P.1 variants of concern V.3. dx.doi.org/10.17504/protocols.io.br9vm966. 2021. (Accessed June 6, 2021, at https://www.protocols.io/view/multiplexed-rt-qpcr-to-screen-for-sars-cov-2-b-1-1-br9vm966.)

16. Abu-Raddad LJ, Chemaitelly H, Butt AA, National Study Group for Covid Vaccination. Effectiveness of the BNT162b2 Covid-19 Vaccine against the B.1.1.7 and B.1.351 Variants. N Engl J Med 2021;385:187–9.

17. Chemaitelly H, Yassine HM, Benslimane FM, et al. mRNA-1273 COVID-19 vaccine effectiveness against the B.1.1.7 and B.1.351 variants and severe COVID-19 disease in Qatar. Nat Med 2021;27:1614–21.

18. Qatar viral genome sequencing data. Data on randomly collected samples. https://www.gisaid.org/phylodynamics/global/nextstrain/. 2021. xat https://www.gisaid.org/phylodynamics/global/nextstrain/.)

19. Benslimane FM, Al Khatib HA, Al-Jamal O, et al. One Year of SARS-CoV-2: Genomic Characterization of COVID-19 Outbreak in Qatar. Front Cell Infect Microbiol 2021;11:768883.

20. Hasan MR, Kalikiri MKR, Mirza F, et al. Real-Time SARS-CoV-2 Genotyping by High-Throughput Multiplex PCR Reveals the Epidemiology of the Variants of Concern in Qatar. Int J Infect Dis 2021;112:52–4.

21. Saththasivam J, El-Malah SS, Gomez TA, et al. COVID-19 (SARS-CoV-2) outbreak monitoring using wastewater-based epidemiology in Qatar. Sci Total Environ 2021;774:145608.

22. El-Malah SS, Saththasivam J, Jabbar KA, et al. Application of human RNase P normalization for the realistic estimation of SARS-CoV-2 viral load in wastewater: A perspective from Qatar wastewater surveillance. Environ Technol Innov 2022;27:102775.

23. Tang P, Hasan MR, Chemaitelly H, et al. BNT162b2 and mRNA-1273 COVID-19 vaccine effectiveness against the SARS-CoV-2 Delta variant in Qatar. Nat Med 2021;27:2136–43.

24. Altarawneh HN, Chemaitelly H, Hasan MR, et al. Protection against the Omicron Variant from Previous SARS-CoV-2 Infection. N Engl J Med 2022;386:1288–90.

25. Chemaitelly H, Ayoub HH, AlMukdad S, et al. Duration of mRNA vaccine protection against SARS-CoV-2 Omicron BA.1 and BA.2 subvariants in Qatar. Nat Commun 2022;13:3082.

26. Qassim SH, Chemaitelly H, Ayoub HH, et al. Effects of BA.1/BA.2 subvariant, vaccination, and prior infection on infectiousness of SARS-CoV-2 omicron infections. J Travel Med 2022.

27. Altarawneh HN, Chemaitelly H, Ayoub HH, et al. Protective Effect of Previous SARS-CoV-2 Infection against Omicron BA.4 and BA.5 Subvariants. N Engl J Med 2022.

28. Chemaitelly H, Tang P, Coyle P, et al. Protection against reinfection with SARS-CoV-2 omicron BA.2.75<sup>*</sup> sublineage. medRxiv 2022:2022.10.29.22281606.

29. World Health Organization. COVID-19 clinical management: living guidance. Available from: https://www.who.int/publications/i/item/WHO-2019-nCoV-clinical-2021-1. Accessed on: May 15, 2021. 2021.

30. World Health Organization. International guidelines for certification and classification (coding) of COVID-19 as cause of death. Available from: https://www.who.int/classifications/icd/Guidelines_Cause_of_Death_COVID-19-20200420-EN.pdf?ua=1. Document Number: WHO/HQ/DDI/DNA/CAT. Accessed on May 15, 2021. 2020.

